# Automated Thoracic Body Composition Analysis on Computed Tomography Is Associated With Ventilator Liberation in Acute Respiratory Failure

**DOI:** 10.64898/2026.07.31.26359447

**Authors:** Pranav Jain, Seyed Mehdi Nouraie, Xin Meng, Tong Yu, Jing Wang, Faraaz Shah, William Bain, Niall Prendergast, Bryan J. McVerry, Anthony Perti, Grant Kokenberger, Jiantao Pu, Georgios D. Kitsios

**Author notes:** Corresponding author: Georgios D. Kitsios, MD, PhD, Assistant Professor of Medicine, Division of Pulmonary, Allergy, Critical Care and Sleep Medicine, University of Pittsburgh Medical Center, Address: UPMC Montefiore Hospital, NW628, 3459 Fifth Avenue, Pittsburgh, PA 15213.

## Abstract

**BACKGROUND:** Liberation from invasive mechanical ventilation (IMV) is a central therapeutic objective in acute respiratory failure (ARF). While lung-protective ventilation strategies and diaphragm function have received considerable attention as determinants of liberation success, the prognostic role of extrapulmonary thoracic tissue composition remains poorly defined. We examined whether chest computed tomography (CT)-derived thoracic skeletal muscle mass (TMM), subcutaneous fat mass (SFM), and their ratio (muscle-fat ratio: MFR) are associated with IMV outcomes in ARF.

**METHODS:** This secondary analysis of a prospective observational ARF cohort includes patients with a chest CT obtained within 7 days before or 3 days after intubation. TMM, SFM and MFR were derived using validated three-dimensional convolutional neural network-based volumetric segmentation spanning the lung apices to bases. Co-primary outcomes were time to successful ventilator liberation within 60 days and 60-day all-cause mortality. Multivariable Cox and logistic regression models with restricted cubic splines assessed linear and non-linear associations, adjusted for age, sex, and key comorbidities. We performed prespecified BMI-stratified and ARDS subgroup analyses.

**RESULTS:** Among 826 patients with ARF requiring IMV, 238 patients with CT imaging were included for analysis (median age 59.5 years, median BMI 28.7 kg/m²). A higher MFR was independently associated with faster ventilator liberation (hazard ratio 1.16, 95% confidence interval [CI] 1.02–1.32, p=0.023). TMM demonstrated a significant J-shaped non-linear relationship with time to liberation (p for non-linearity=0.019), with delayed liberation at both low and high extremes of TMM. Higher SFM was independently associated with increased 60-day mortality (odds ratio 1.09, 95% CI 1.00–1.18, p=0.049). In 81 patients with ARDS, TMM also demonstrated a significant J-shaped non-linear relationship with 60-day mortality (p for non-linearity=0.037). BMI was not significantly associated with either outcome.

**CONCLUSIONS:** Volumetric CT-derived thoracic body composition metrics, particularly MFR and TMM, are independently associated with ventilator liberation and 60-day mortality in ARF, capturing prognostic information not reflected by BMI alone. These findings support the incorporation of automated thoracic body composition analysis into early risk stratification frameworks for mechanically ventilated patients and highlight extrapulmonary tissue composition as an underexplored determinant of IMV outcomes.

**Key Points:** *Question:* In adults with acute respiratory failure on invasive mechanical ventilation, are CT-derived thoracic skeletal muscle mass (TMM), subcutaneous fat mass (SFM), and muscle-fat ratio (MFR) associated with ventilator liberation and 60-day mortality?

*Findings:* In this analysis of a prospective cohort of 238 patients, higher MFR was associated with faster liberation and TMM showed a J-shaped association with liberation. Higher SFM predicted greater 60-day mortality, while BMI category predicted neither.

*Meaning:* Automated CT-derived thoracic body composition adds prognostic information beyond BMI and may aid early risk stratification in mechanically ventilated ARF patients.

## Introduction

Liberation from invasive mechanical ventilation (IMV) is a central therapeutic objective in the management of acute respiratory failure (ARF). While IMV is lifesaving, prolonged mechanical ventilation (PMV) carries substantial independent risks, including ventilator-induced lung injury, diaphragmatic dysfunction, skeletal muscle weakness, delirium, and increased susceptibility to secondary infections ^1,2^. More than one-third of patients who reach readiness for spontaneous breathing trials attempts experience difficult or prolonged weaning ^3,4^. Those requiring PMV face markedly higher mortality, long-term neurocognitive impairment, and reduced quality of life compared to patients liberated early ^2^. The healthcare utilization and costs associated with PMV place a substantial burden on patients, families, and health systems alike ^2,5,6^.

Two domains have appropriately received considerable attention as determinants of liberation success. First, lung-protective ventilation strategies including tidal volume and driving pressure limitation, combined with individualized positive end-expiratory pressure (PEEP) titration, have established a foundation for safe IMV management ^7–9^. Second, low diaphragm muscle mass and reduced thickening fraction, as assessed by bedside ultrasound or CT, are independently associated with prolonged ventilation and higher mortality ^10,11^. Strategies to detect and treat diaphragm dysfunction, including inspiratory muscle training and temporary transvenous phrenic nerve stimulation, are accordingly active areas of investigation ^12^.

By contrast, the role of extrapulmonary thoracic tissue composition in determining IMV outcomes has received comparatively little systematic study. The thoracic musculature and subcutaneous adipose tissue represent substantial compartments visible on chest CT scans routinely obtained in ARF. These tissues have direct physiological relevance to respiratory mechanics. Increased chest wall mass reduces total respiratory system compliance without altering intrinsic lung compliance. This mechanical consequence is clinically significant in patients managed with plateau pressure targets, as increased chest wall load may necessitate higher driving pressures and complicate ventilator liberation independent of underlying lung pathology. Yet the relationship between thoracic tissue composition and ventilator liberation and clinical outcomes remains poorly defined.

This gap is particularly relevant given that body mass index (BMI), the most commonly used somatometric measure in critical illness, conflates lean muscle mass and adipose tissue, obscuring clinically meaningful differences in somatic reserve – the body’s nutritional and functional capacity to sustain the metabolic and mechanical demands of weaning. BMI-based analyses in ARF have produced inconsistent results and do not capture the relative contributions of muscle versus fat that may independently influence weaning capacity ^13–17^.

Automated CT-based body composition offers a more granular approach. Skeletal muscle mass and subcutaneous fat mass can be derived from CT imaging using validated convolutional neural networks (CNN). Preliminary data in patients with chronic obstructive pulmonary disease (COPD), COVID-19, and acute respiratory distress syndrome (ARDS) suggest that these metrics carry prognostic information ^18–21^. In mechanically ventilated patients with hematologic malignancies, a higher fat-to-muscle cross-sectional area (CSA) ratio at T12 was associated with longer time to ventilator liberation^22^. However, prior studies have used heterogeneous methods, including CSA measurements at a single vertebral level, limiting generalizability. A volumetric approach quantifying actual tissue mass across the entire thorax may offer more comprehensive and biologically relevant estimates.

In this analysis of a prospectively enrolled ARF cohort, we examined whether CT-derived thoracic skeletal muscle mass (TMM), thoracic subcutaneous fat mass (SFM), and their ratio (MFR = TMM/SFM) are associated with time to successful liberation from IMV and 60-day mortality in patients with ARF. We hypothesized that a higher MFR would be associated with faster ventilator liberation and lower mortality, reflecting a more favorable somatic reserve profile at ICU admission.

## Materials and Methods

### Study design and setting

This is an analysis of the Pittsburgh Acute Lung Injury Registry and Biospecimen Repository (ALIR), a prospective observational cohort of patients admitted to ICUs at UPMC hospitals between 2011 and 2024. ALIR prospectively captures detailed demographic, clinical, laboratory, biomarker, and outcome data on enrolled participants. Patients were eligible for the present analysis if they required endotracheal intubation and IMV for ARF and had at least one chest CT scan obtained within 7 days before or 3 days after intubation. This peri-intubation window was chosen because thoracic tissue composition at this early time point is unlikely to have been substantially altered by the acute illness, and therefore reflects premorbid nutritional and functional reserve rather than catabolic changes accrued during the ICU course. Although brief exposure to mechanical ventilation can initiate skeletal muscle catabolism, the post-intubation limit of 3 days was selected as a pragmatic threshold to capture tissue composition that is still predominantly reflective of the patient’s premorbid state ^23^.

### ARF Subtype Classification

Participants were systematically classified into one of seven predefined ARF subtypes based on clinical data, chest imaging, and laboratory results: ARDS (Berlin definition), at-risk for ARDS (ARFA; identifiable ARDS risk factor without fulfilling ARDS criteria), congestive heart failure–related cardiogenic pulmonary edema, airway control without primary pulmonary pathology, acute-on-chronic hypercapnic respiratory failure from obstructive airways disease, acute exacerbation of interstitial lung disease, and other/multifactorial when features overlapped or did not clearly fit a single category ^24^. Classification was performed by a consensus committee of board-certified critical care physicians blinded to research biomarkers and outcomes.

### Cohort assembly

Of 826 ALIR participants with ARF requiring IMV, 366 had chest CT scans technically adequate for CNN–based body composition analysis. Of these, 258 scans were acquired within the peri-intubation window. When multiple eligible CT scans existed, the scan closest to the date of intubation was selected; pre-intubation scans were preferred when two scans were equidistant. This yielded 239 unique patients, of whom 238 comprised the final ARF analytic cohort after excluding one patient for missing data. Characteristics of those included in the analysis with CT within peri-intubation window and those without are compared in Supplementary Table 1. Patients meeting Berlin criteria for ARDS were identified as a prespecified subgroup (n=81).

### CT-derived body composition metrics

Thoracic body composition was quantified from chest CT scans using previously developed and validated three-dimensional CNN in collaboration with the Departments of Radiology and Bioengineering at the University of Pittsburgh ^25^. Automated volumetric segmentation spanned the lung apices to bases (Figure 1), explicitly excluding the diaphragm from the skeletal muscle compartment. Three exposure variables were defined for each patient: thoracic skeletal muscle mass (TMM, kg), thoracic subcutaneous fat mass (SFM, kg), and their ratio (MFR = TMM/SFM).

**Figure 1.**
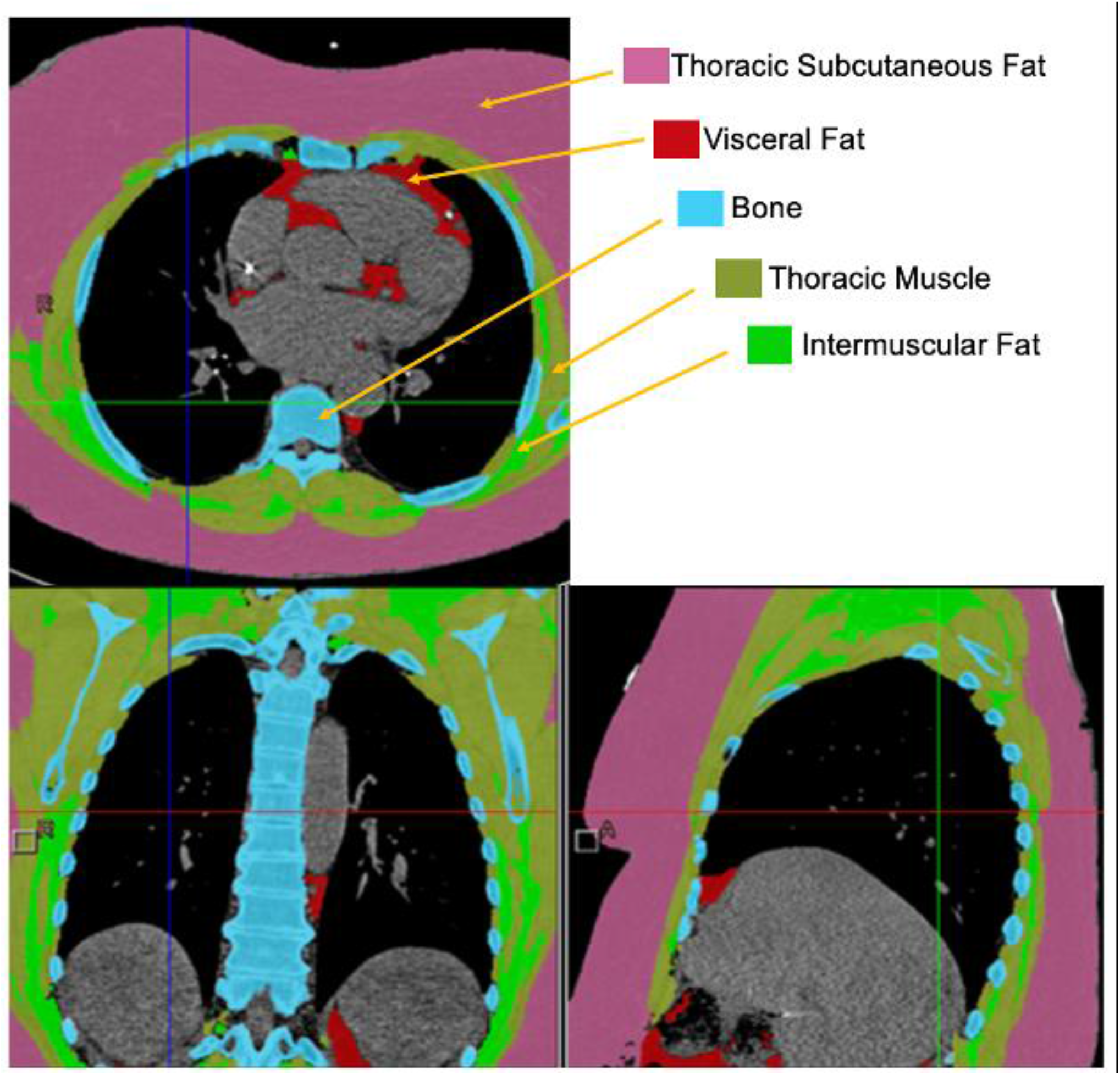
Automated Segmentation of Thoracic Body Composition Metrics in Patients with Acute Respiratory Failure. Axial, coronnal, and sagital thoracic computed tomography images with demonstration of automated segmentation of body tissues. Based on the density and volume of tissue, the mass of thoracic muscle and subcutaneous fat as measured from the apex to the base of the lung for each patient was obtained.

CT-derived lung parenchymal metrics: Total lung volume and volume of consolidated lung for each patient were also derived from the chest CT scans using the described software.

### Outcomes

The co-primary outcomes were time to successful ventilator liberation within 60 days of intubation and 60-day all-cause mortality. Time to successful ventilator liberation was defined as liberation from IMV within 60 days without death occurring during that same period, consistent with contemporary ventilator-free day endpoint methodology ^26^. A key secondary outcome was clinical trajectory, with three mutually exclusive groups defined as: “Early Liberation” (successful extubation with <14 days of IMV and survival to 60 days), “PMV” (successful extubation with ≥14 days IMV and survival to 60 days) ^1^, and “Non-Survivors” (death within 60 days).

### Statistical analysis

Baseline characteristics are reported as means (SD), medians (IQR), or counts (%) as appropriate. Spearman rank correlation coefficients were used to assess bivariate associations between CT-derived body composition metrics (TMM, SFM, and MFR) and thoracic volumetric variables (total lung volume and consolidative lung volume) with Day 1 respiratory mechanics (respiratory compliance, normalized respiratory elastance, and average PEEP) given the non-normal distribution of these variables.

Cox proportional hazards regression was used to analyze time to ventilator liberation for each of TMM, SFM, and MFR in the ARF cohort; hazard ratios (HR) >1 indicate faster liberation. Proportional hazards (PH) assumption was tested using scaled Schoenfeld residuals over time. In all models, PH assumption held for the primary covariate (p-value > 0.05). Multivariable logistic regression was used to estimate associations with 60-day mortality. Covariates for multivariable models were selected based on a directed acyclic graph (DAG, *Supplementary Figure 1*) and constrained to a maximum of 8 total variables to maintain an events-per-variable (EPV) ratio appropriate to the number of mortality outcomes observed in the ARF cohort. Congestive heart failure, active neoplasm, and chronic liver disease were identified a priori on the DAG but excluded from the final model due to their lower prevalence within the cohort. All models were adjusted for age and sex. Sex was included to account for body composition differences related to breast tissue; BMI was excluded to avoid collinearity with CT-derived metrics. Restricted cubic splines (RCS) with three knots at the 10th, 50th, and 90th percentiles of each exposure were incorporated into Cox and logistic models to evaluate non-linearity, with joint tests of spline terms used to assess departure from linearity. As an additional exploratory analysis, Wilcoxon rank-sum testing was performed to compare TMM between patients who received neuromuscular blocking agents (NMBA) during their ICU stay following intubation and those who did not. This was done given the potential for NMBA use to reflect illness severity and the hypothesis that patients with greater TMM may generate higher respiratory effort and greater patient-ventilator dyssynchrony, thereby necessitating NMBA use and contributing to PMV.

To characterize clinical trajectories, multinomial logistic regression with Early Liberation as the reference group was used to estimate associations between each CT-derived body composition metric and membership in the PMV or Non-Survivor trajectory groups, with all models adjusted for age and sex. Differences in body composition metrics across the three clinical trajectory groups were assessed using Kruskal-Wallis tests with pairwise comparisons performed using the Dunn test. P-values for all pairwise comparisons were adjusted for multiple testing using the Bonferroni correction.

As secondary analyses, all primary regression models were repeated in the prespecified ARDS subgroup. Given the limited number of mortality outcomes in this subgroup, multivariable models were adjusted only for age to preserve an adequate EPV ratio. BMI was also tested as a primary covariate in 60 day time to ventilator liberation and 60 day mortality models. Outcomes were additionally compared across four BMI strata (normal weight ≥18.5–<25, overweight ≥25–<30, obesity class I ≥30–<35, and obesity class II/III ≥35 kg/m²) among the 231 ARF patients with BMI ≥18.5 kg/m² using chi-square tests for categorical outcomes and Kruskal-Wallis tests for continuous variables. Because BMI conflates lean and adipose tissue mass into a single scalar measure, these analyses were intended to evaluate whether a crude somatometric index captures prognostic information in ARF and to contextualize the incremental value of tissue-specific CT-derived metrics over conventional body habitus classification. All tests were two-sided with a significance threshold of α=0.05. Statistical analyses were performed using R statistical software, version 4.5.3 (R Foundation for Statistical Computing, Vienna, Austria).

### Ethics

The ALIR protocol and the specific analyses in this study were approved by the University of Pittsburgh IRB (STUDY19050099). Informed consent was obtained from participants or their legally authorized representatives at the time of enrollment. All procedures followed the ethical standards of the Declaration of Helsinki.

### Use of Generative AI

Claude Opus models were utilized for improving spelling, language, grammar, and syntax of manuscript text and figure/table legends and footnotes. Claude Opus models were also used to aid in generating R code for statistical analysis. The author thoroughly reviewed all output for accuracy.

## Results

Detailed demographics, comorbidities, illness severity, and CT-derived body composition metrics for the full cohort of 238 patients and the ARDS subgroup are presented in *Table 1*. The cohort median age was 59.5 (IQR 44.0 – 68.8) years and the median BMI was 28.7 (IQR 24.6 – 35.0) kg/m². ARF subtype distribution is shown in Supplementary *Figure 2*. The 81-patient ARDS subgroup was younger (median age 55.3, IQR 38.7 – 63.9 years) with a similar BMI (median 29.0, IQR 24.2 – 35.2 kg/m²). Median TMM, SFM, and MFR for both the overall ARF cohort and the ARDS subgroup are displayed in *Table 1*. Patients were classified into three mutually exclusive 60-day trajectory groups: Early Liberation (n=127, 53.4%), PMV (n=28, 11.8%), and Non-Survivors (n=83, 34.9%). Group distribution and baseline characteristics, including TMM, SFM, and MFR across trajectory groups, are presented in *Table 2*.

**Table 1.** Baseline Characteristics of ARF and ARDS Patients.

|  | All ARF | ARDS |
| --- | --- | --- |
| Characteristic | All ARF N = 238<sup>1</sup> | ARDS N = 81<sup>1</sup> |
| **Age (years)** | 59.47 (48.00 - 67.77) | 55.31 (38.72 - 63.87) |
| **Sex** |  |  |
| Male | 127 (53.4%) | 37 (45.7%) |
| Female | 111 (46.6%) | 44 (54.3%) |
| **Body Mass Index (kg/m<sup>2</sup>)** | 28.65 (24.59 - 34.96) | 28.96 (24.16 - 35.18) |
| **Thoracic Muscle Mass (kg)** | 3.50 (2.95 - 4.71) | 3.36 (2.92 - 4.47) |
| **Subcutaneous Fat Mass (kg)** | 3.76 (2.48 - 5.88) | 3.72 (2.58 - 5.76) |
| **Muscle-to-Fat Ratio** | 0.96 (0.64 - 1.40) | 1.00 (0.64 - 1.33) |
| **History of Alcohol Use** | 36 (15.1%) | 8 (9.9%) |
| **History of Congestive Heart Failure** | 27 (11.3%) | 5 (6.2%) |
| **History of Chronic Liver Disease** | 14 (5.9%) | 4 (4.9%) |
| **History of COPD** | 54 (22.7%) | 12 (14.8%) |
| **History of Chronic Renal Failure** | 51 (21.4%) | 15 (18.5%) |
| **History of Diabetes** | 76 (31.9%) | 21 (25.9%) |
| **Active Neoplasm** | 14 (5.9%) | 2 (2.5%) |
| **Vasopressors on Intubation** | 155 (65.1%) | 51 (63.0%) |
| **History of Hypoalbuminemia** | 149 (62.6%) | 49 (60.5%) |
| **History of Pulmonary Fibrosis** | 16 (6.7%) | 7 (8.6%) |
| **Hemodynamic Instability on Intubation** | 11 (4.6%) | 2 (2.5%) |
| **History of Immunosuppression** | 55 (23.1%) | 25 (30.9%) |
| **60-Day Mortality** | 83 (34.9%) | 23 (28.4%) |
| **60-Day Successful Liberation** | 154 (64.7%) | 57 (70.4%) |

<sup>1</sup>Continuous variables presented as median (P25-P75); categorical variables as n (%).

**Table 2.** Baseline Characteristics by Clinical Trajectory Group.

| Characteristic | Clinical Trajectory Group |  |  | P-value<sup>2</sup> |
| --- | --- | --- | --- | --- |
|  | Early Liberation N = 127<sup>1</sup> | Prolonged MV N = 28<sup>1</sup> | Non-Survivor N = 83<sup>1</sup> |  |
| **Age (years)** | 56.74 (44.97 - 66.87) | 51.71 (36.79 - 62.41) | 64.71 (55.39 - 72.15) | **<0.001** |
| **Sex** |  |  |  | 0.596 |
| Male | 65 (51.2%) | 14 (50.0%) | 48 (57.8%) |  |
| Female | 62 (48.8%) | 14 (50.0%) | 35 (42.2%) |  |
| **Body Mass Index (kg/m<sup>2</sup>)** | 29.75 (23.66 - 35.13) | 28.05 (23.75 - 36.42) | 28.34 (25.15 - 34.62) | 0.879 |
| **Thoracic Muscle Mass (kg)** | 3.55 (2.99 - 4.79) | 3.33 (2.56 - 4.24) | 3.46 (2.86 - 4.74) | 0.348 |
| **Subcutaneous Fat Mass (kg)** | 3.67 (2.34 - 5.71) | 3.69 (2.05 - 5.30) | 4.08 (2.71 - 7.03) | 0.195 |
| **Muscle-to-Fat Ratio** | 1.03 (0.62 - 1.54) | 0.92 (0.65 - 1.42) | 0.86 (0.64 - 1.20) | 0.276 |
| **History of Alcohol Use** | 18 (14.2%) | 1 (3.6%) | 17 (20.5%) | 0.088 |
| **History of Congestive Heart Failure** | 12 (9.4%) | 0 (0.0%) | 15 (18.1%) | **0.021** |
| **History of Chronic Liver Disease** | 4 (3.1%) | 1 (3.6%) | 9 (10.8%) | 0.059 |
| **History of COPD** | 28 (22.0%) | 0 (0.0%) | 26 (31.3%) | **0.003** |
| **History of Chronic Renal Failure** | 26 (20.5%) | 4 (14.3%) | 21 (25.3%) | 0.437 |
| **History of Diabetes** | 50 (39.4%) | 8 (28.6%) | 18 (21.7%) | **0.025** |
| **Active Neoplasm** | 7 (5.5%) | 0 (0.0%) | 7 (8.4%) | 0.252 |
| **Vasopressors on Intubation** | 79 (62.2%) | 15 (53.6%) | 61 (73.5%) | 0.096 |
| **History of Hypoalbuminemia** | 77 (60.6%) | 19 (67.9%) | 53 (63.9%) | 0.742 |
| **History of Pulmonary Fibrosis** | 5 (3.9%) | 4 (14.3%) | 7 (8.4%) | 0.105 |
| **Hemodynamic Instability on Intubation** | 7 (5.5%) | 0 (0.0%) | 4 (4.8%) | 0.451 |
| **History of Immunosuppression** | 27 (21.3%) | 9 (32.1%) | 19 (22.9%) | 0.465 |
| **60-Day Mortality** | 0 (0.0%) | 0 (0.0%) | 83 (100.0%) | **<0.001** |
| **60-Day Successful Liberation** | 127 (100.0%) | 27 (96.4%) | 0 (0.0%) | **<0.001** |

<sup>1</sup>Continuous variables presented as median (P25-P75); categorical variables as n (%). P-values from Kruskal-Wallis test for continuous variables and Chi-square test for categorical variables.
<sup>2</sup>Kruskal-Wallis rank sum test; Pearson's Chi-squared test

### Thoracic Body Composition, Lung Parenchymal Metrics, and Baseline Respiratory Mechanics

Respiratory system compliance and elastance measured on the first day of IMV integrate contributions from both the lung parenchyma and the extrapulmonary thoracic compartment including the chest wall, thoracic musculature, and adipose tissue. To characterize the independent relationships of each compartment with respiratory mechanics on the first day of IMV, we examined cross-sectional correlations of TMM, SFM, and MFR alongside CT-derived parenchymal metrics of total lung volume and consolidated lung volume with respiratory system compliance, normalized elastance, and applied PEEP in 234 patients with complete data. CT body composition metrics at best showed weak correlations with respiratory mechanics. TMM was weakly but significantly correlated with respiratory system compliance (ρ=0.275, p<0.001) and applied PEEP (ρ=0.189, p=0.004), but was not significantly correlated with normalized elastance (ρ=−0.018, p=0.788; *Supplementary Figure 3*). SFM similarly showed at most weak correlations, while MFR showed negligible correlations (*Supplementary Figures 4 and 5*). Among the parenchymal metrics, total lung volume was weakly but positively correlated with compliance (ρ=0.302, p<0.001). Lung volume and consolidation volume otherwise showed weak correlations with respiratory mechanics (*Supplementary Figures 6 and 7*).

### Time to successful ventilator liberation within 60 days

In the ARF cohort, TMM was not linearly associated with time to successful ventilator liberation within 60 days (HR 0.96, 95% CI 0.87–1.05, p=0.357); however, RCS demonstrated a significant non-linear component for TMM (p for non-linearity=0.019), with a J-shaped pattern suggesting higher risk of delayed liberation at both low and high extremes of TMM. Higher SFM demonstrated a trend toward slower liberation that did not reach statistical significance (HR 0.95, 95% CI 0.91–1.00, p=0.067), and the non-linear component was not significant (p=0.887). A higher MFR was significantly associated with faster successful ventilator liberation (HR 1.16, 95% CI 1.02–1.32, p=0.023). RCS analysis did not indicate evidence of non-linearity for this association (p=0.308). History of diabetes mellitus (DM) was independently associated with faster time to ventilator liberation across all three ARF time-to-event models, while older age was associated with slower liberation in all models. Results are displayed in *Figure 2* and *Supplementary Tables 2 and 3*.

**Figure 2.**
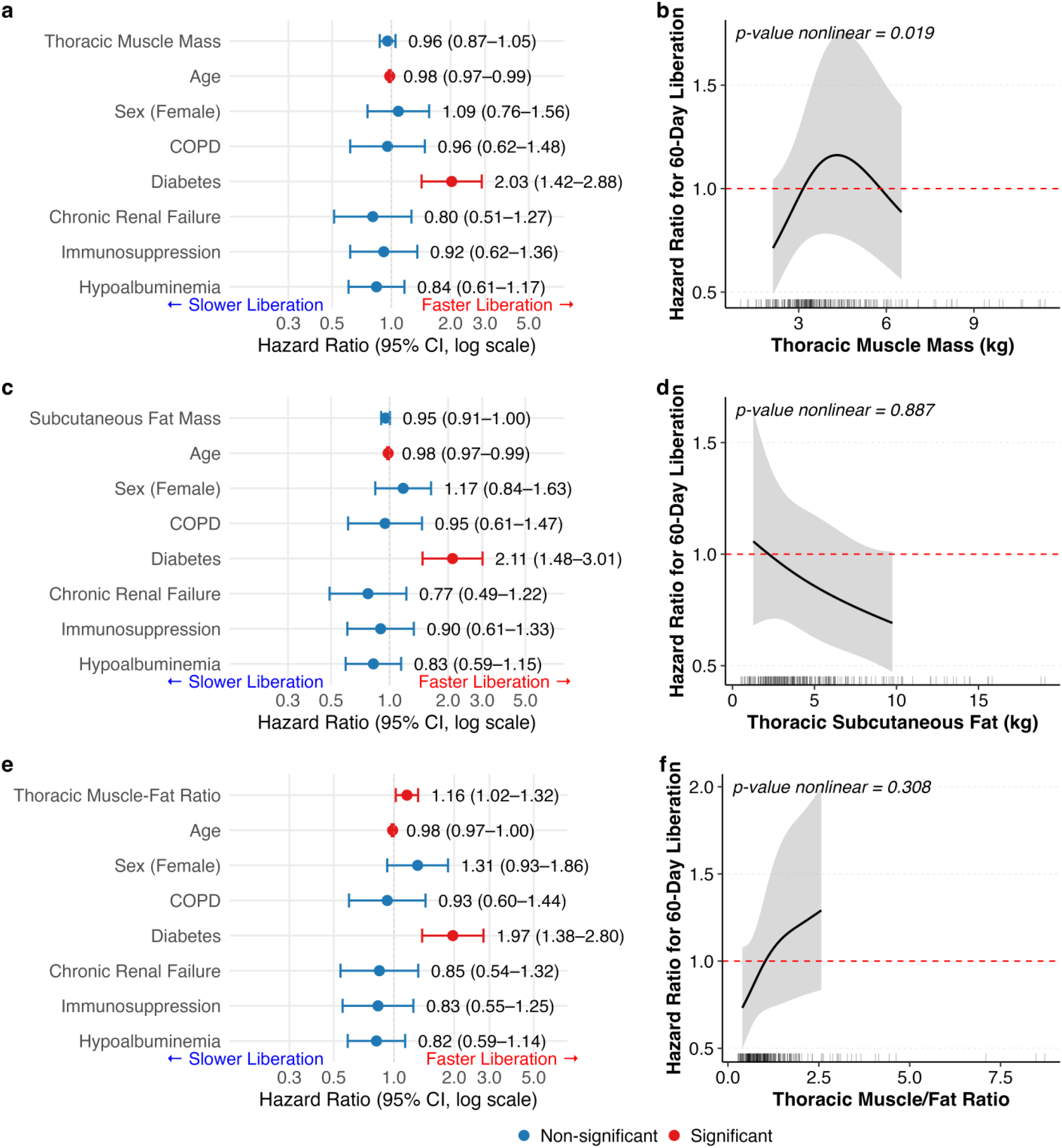
Association Between Thoracic Body Composition Metrics and Time to Successful Ventilator Liberation within 60 Days in Patients with Acute Respiratory Failure. Panels (a), (c), and (e) display forest plots from multivariable Cox proportional hazards regression models examining the association of thoracic muscle mass (a), thoracic subcutaneous fat mass (c), and thoracic muscle-to-fat ratio (e) with 60-day ventilator liberation, adjusted for 7 covariates as displayed in ARF. Each point represents the hazard ratio (HR) with 95% confidence intervals (CI); values greater than 1 indicate faster liberation and values less than 1 indicate slower liberation. Red points and error bars denote statistically significant associations (p < 0.05); blue denotes non-significant associations. Panels (b), (d), and (f) display restricted cubic spline (RCS) curves from corresponding Cox regression models, illustrating the dose-response relationship between each body composition metric and the hazard of liberation. The shaded region represents the 95% CI. The dashed red horizontal line indicates a hazard ratio of 1 (no effect). Rug marks along the x-axis reflect the distribution of observed values. The p-value for the nonlinear component of each spline is shown in the upper left corner of each RCS panel. Abbreviations: ARF, acute respiratory failure;HR, hazard ratio;CI, confidence interval;RCS, restricted cubic spline;COPD, chronic obstructive pulmonary disease.

### 60-day mortality: multivariable logistic regression

In the ARF cohort, TMM showed a non-significant association with 60-day mortality on linear analysis (OR 1.12, 95% CI 0.95–1.32, p=0.185). RCS analysis revealed a significant non-linear relationship between TMM and 60-day mortality (p for non-linearity=0.021), with the estimated relationship appearing J-shaped across the exposure distribution. SFM was significantly associated with higher 60-day mortality risk (OR 1.09, 95% CI 1.00–1.18, p=0.049), and RCS terms did not suggest deviation from linearity (p for non-linearity=0.978). Though not reaching statistical significance, higher MFR did trend towards reduced 60-day mortality (OR 0.66, 95% CI 0.43–1.02, p=0.066), and the non-linear component was similarly non-significant (p for non-linearity=0.517). History of DM was independently associated with decreased odds of 60-day mortality across all three ARF models, while older age was significantly associated with higher odds of 60-day mortality in all three models. Results are displayed in *Figure 3* and *Supplementary Tables 4 and 5*.

**Figure 3.**
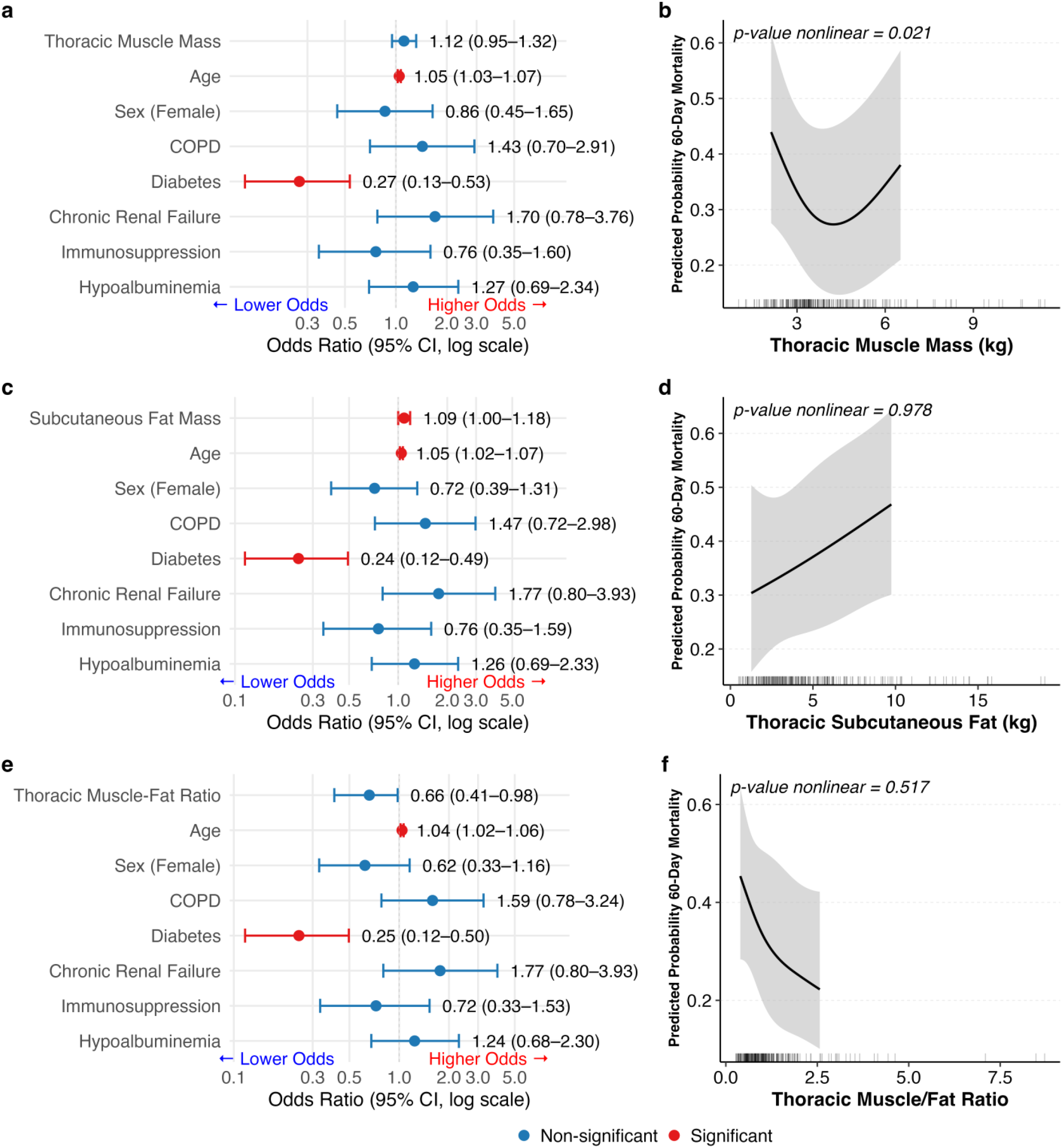
Association Between Thoracic Body Composition Metrics and 60-Day Mortality in Patients with Acute Respiratory Failure. Panels (a), (c), and (e) display forest plots from multivariable logistic regression models examining the association of thoracic muscle mass (a), thoracic subcutaneous fat mass (c), and thoracic muscle-to-fat ratio (e) with 60-day mortality, adjusted for 7 covariates as displayed in ARF. Each point represents the odds ratio (OR) with 95% confidence intervals (CI); values greater than 1 indicate increased odds of mortality (risk factor) and values less than 1 indicate decreased odds of mortality (protective). Red points and error bars denote statistically significant associations (p < 0.05); blue denotes non-significant associations. Panels (b), (d), and (f) display restricted cubic spline (RCS) curves from corresponding logistic regression models, illustrating the dose-response relationship between each body composition metric and the odds of 60-day mortality. The shaded region represents the 95% CI. The dashed red horizontal line indicates an odds ratio of 1 (no effect). Rug marks along the x-axis reflect the distribution of observed values. The p-value for the nonlinear component of each spline is shown in the upper left corner of each RCS panel. Abbreviations: ARF, acute respiratory failure; OR, odds ratio; CI, confidence interval; RCS, restricted cubic spline; COPD, chronic obstructive pulmonary disease.

### Clinical Trajectory Group Analysis

To evaluate whether body composition metrics differentiated patients across 60-day clinical trajectories, multinomial logistic regression was performed with Early Liberation as the reference category, adjusting for age and sex. Higher TMM showed a trend toward lower odds of the PMV outcome relative to Early Liberation (OR 0.72, 95% CI 0.51–1.03, p=0.074) but was not significantly associated with the Non-Survivor outcome (OR 1.09, 95% CI 0.93–1.28, p=0.299). SFM and MFR were also not significantly associated with either the PMV or Non-Survivor trajectory relative to Early Liberation (Supplementary Table 6). Kruskal-Wallis testing revealed no statistically significant difference in TMM (H=2.11, p=0.348), SFM (H=3.27, p=0.195), or MFR (H=2.58, p=0.276) across the three trajectory groups. Dunn pairwise post-hoc comparisons with Bonferroni correction likewise demonstrated no significant pairwise differences between Early Liberation, PMV, and Non-Survivor groups for any body composition metric (*Figure 4*).

**Figure 4.**
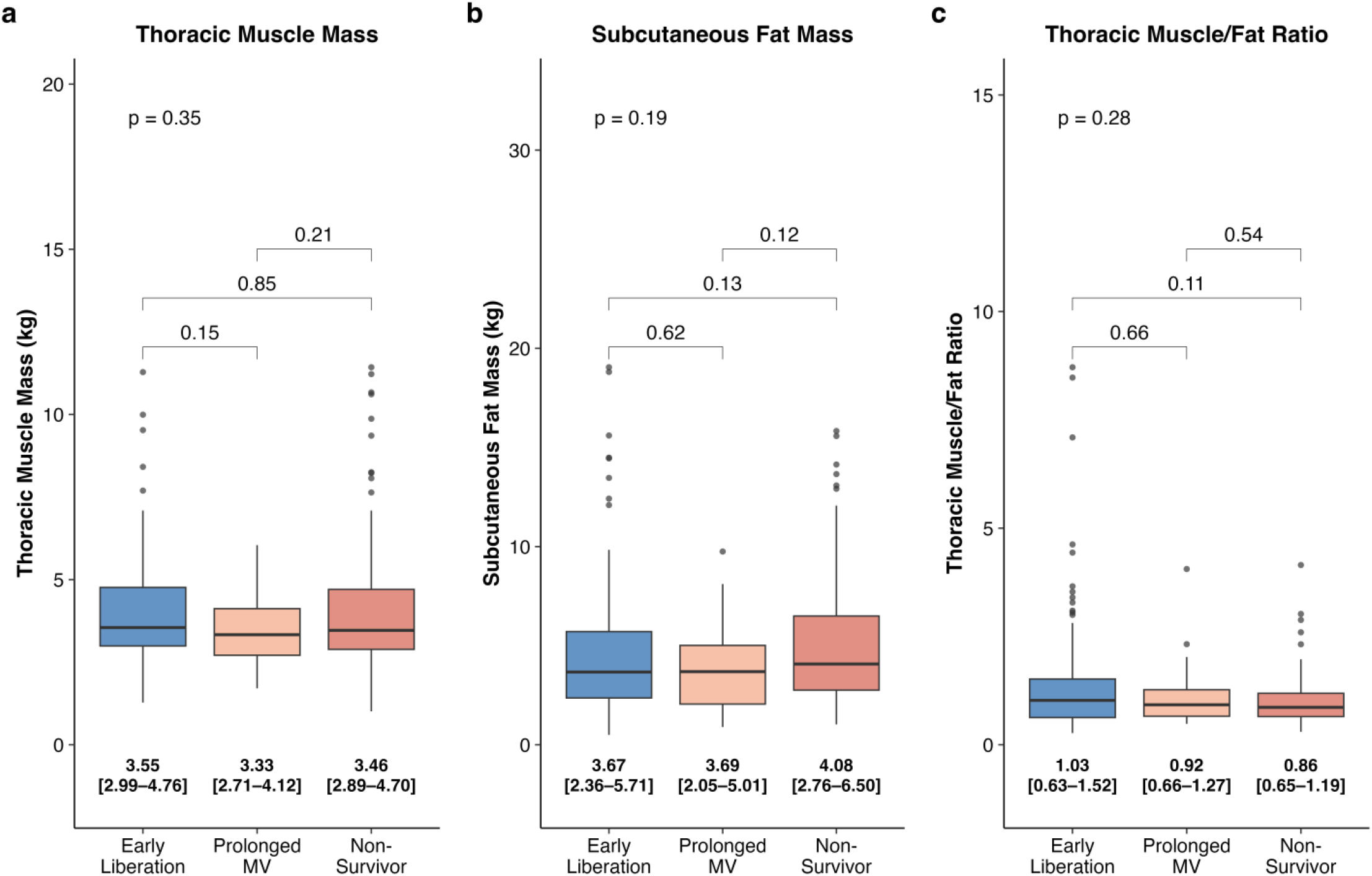
CT-Derived Thoracic Body Composition Metrics Across 60-Day Clinical Trajectory Groups. Distribution of (a) TMM (kg), (b) SFM (kg), and (c) MFR across three mutually exclusive 60-day clinical trajectory groups: Early Liberation (successful extubation ≤14 days with survival to 60 days), Prolonged Mechanical Ventilation (successful extubation >14 days with survival to 60 days), and Non-Survivors (death within 60 days). Box plots display median (center line), interquartile range (box), and 1.5×IQR (whiskers); values in bold below each box represent median [Q1–Q3]. Overall group differences were assessed by Kruskal-Wallis test; pairwise comparisons were performed using Wilcoxon rank-sum testing with Bonferroni correction for multiple comparisons. P-values are displayed above each pairwise bracket and the overall Kruskal-Wallis p-value is shown above all brackets. Abbreviations: CT, computed tomography; TMM, thoracic skeletal muscle mass; SFM, thoracic subcutaneous fat mass; MFR, muscle-to-fat ratio; MV, mechanical ventilation; IQR, interquartile range.

### Neuromuscular Blockade Analysis

Among the full ARF cohort, 44 (18.5%) patients received NMBA during their ICU stay post intubation (ie. not inclusive of single NMBA doses used for intubation). TMM did not differ significantly between patients who received NMBA and those who did not (median difference −0.16 kg, 95% CI −0.65 to 0.39; Wilcoxon rank-sum p=0.584). Results are displayed in *Supplementary Figure 8*.

### ARDS subgroup analyses

In the ARDS subgroup (n=81), none of the body composition metrics were significantly associated with time to successful ventilator liberation within 60 days. The HR for TMM was 0.85 (95% CI 0.72–1.02, p=0.077), for SFM was 0.92 (95% CI 0.83–1.03, p=0.141), and for MFR was 1.05 (95% CI 0.85–1.30, p=0.629). RCS analysis did not demonstrate a statistically significant non-linear component for TMM in this subgroup (p for non-linearity=0.094), though a trend was observed; spline analysis similarly did not indicate evidence of non-linearity for SFM or MFR (p=0.273 and p=0.855, respectively). Older age was significantly associated with lower probability of successful ventilator liberation across all three ARDS models. Results are displayed in *Supplementary Figure 9* and *Supplementary Tables 7 and 8*.

TMM in ARDS was significantly associated with higher odds of 60-day mortality on linear analysis (OR 1.45, 95% CI 1.03–2.07, p=0.036), a notable divergence from the overall ARF cohort findings. RCS analysis confirmed a significant non-linear relationship between TMM and 60-day mortality in this subgroup (p for non-linearity=0.037), with the predicted probability curve appearing J-shaped. SFM demonstrated a non-significant trend toward higher 60-day mortality (OR 1.19, 95% CI 0.97–1.46, p=0.085), and RCS analysis did not demonstrate significant non-linearity (p for non-linearity=0.213). MFR was not significantly associated with 60-day mortality in the ARDS subgroup (OR 0.68, 95% CI 0.27–1.72, p=0.420), with no evidence of non-linearity on spline analysis (p for non-linearity=0.312). Older age remained a significant predictor of 60-day mortality across all three ARDS subgroup models. Results are displayed in *Supplementary Figure 10* and *Supplementary Tables 9 and 10*.

### BMI analyses

BMI demonstrated no association with 60 day time to liberation (HR 1.00, 95% CI 0.98-1.02, p=0.803) or 60 day mortality (OR 1.01, 95% CI 0.98-1.04, p=0.589). BMI RCS models of time to liberation (p for non-linearity=0.749) and mortality (p for non-linearity=0.747) are without evidence of non-linearity. Results are displayed in *Supplementary Figure 11 and Supplmentary Table 11*. In analyses restricted to ARF patients with BMI ≥18.5 kg/m² (n=231), there were no statistically significant differences in 60-day mortality across normal weight, overweight, obesity class I, and obesity class II/III strata (global chi-square p=0.432). Similarly, the proportion of patients achieving successful ventilator liberation within 60 days did not differ significantly across BMI groups (global chi-square p=0.327). TMM (p=0.002), SFM (p= < 0.001), and MFR (p= < 0.001) differed across BMI strata as assessed by Kruskal-Wallis tests. Age also differed across BMI strata (p = 0.036). Results are displayed in *Supplementary Table 12*.

## Discussion

This prospective cohort study with secondary analysis of CT imaging demonstrates that CT-derived thoracic body composition metrics are associated with successful liberation from IMV and 60-day mortality in ARF, whereas BMI was not meaningfully related to short-term outcomes. A higher MFR was associated with faster time to successful ventilator liberation within 60 days. TMM demonstrated a significant J-shaped non-linear relationship with both time to liberation and 60-day mortality, with both low and high extremes of TMM associated with worse outcomes. SFM was significantly associated with higher 60-day mortality on linear analysis without evidence of non-linearity. ARDS subgroup analyses for time to liberation were likely underpowered to detect meaningful differences, though TMM was significantly associated with 60-day mortality in this subgroup with a similarly J-shaped non-linear pattern. The clinical trajectory analysis classifying patients as Early Liberation, PMV, or Non-Survivors did not demonstrate statistically significant differences in TMM, SFM, or MFR across groups on either Kruskal-Wallis testing or multinomial logistic regression. The lack of association seen may reflect loss of statistical power due to converting the continuous time-to-event data into the described threshold-defined categories as well as heterogeneity within trajectory groups, especially amongst non-survivors who died at varying timepoints during the ICU course. The trajectory analysis should therefore be interpreted as an exploratory characterization of the cohort rather than used to refute the associations observed in the primary regression analyses.

A key methodological innovation of this study is the quantification of thoracic tissue mass (kg) from the lung apices to the bases which the full three-dimensional volumetric extent of both the thoracic skeletal musculature and subcutaneous fat compartment. Prior studies in this space have primarily utilized CSA measurements at single fixed anatomic landmark such as the level of the carina or a specific thoracic vertebral body. CSA methods sample only a thin axial slice of a heterogeneous three-dimensional tissue which may not fully reflect the global somatic reserve that influences respiratory mechanics and recovery capacity. By contrast, the volumetric CNN-based segmentation used in this study integrates tissue mass across the entire thorax providing estimates that are more biologically representative of the total musculoskeletal and adipose reservoir. It is important to note that our approach explicitly excluded the diaphragm and thus the TMM metric only reflects accessory and thoracic cage musculature.

CT-derived thoracic body composition metrics showed overall weak correlations with respiratory system mechanics on Day 1 of IMV. TMM was weakly but significantly correlated with respiratory system compliance and applied PEEP while SFM showed similarly weak correlations and MFR showed negligible relationships with respiratory mechanics. These findings suggest that thoracic body composition as measured by volumetric segmentation of the musculature and subcutaneous fat compartment while excluding the diaphragm may play a relatively limited direct role in determining acute respiratory mechanics at the time of intubation. Instead, the prognostic associations between body composition metrics and liberation and mortality observed in this study may be more reflective of factors such as systemic somatic reserve and the body’s inflammatory and metabolic response to the physiologic insult of ARF requiring IMV. Low muscle mass has been linked to impaired protein synthesis, altered cytokine responses, and higher infection risk ^27–30^, while excess adipose tissue may promote a pro-inflammatory, insulin-resistant state that impairs weaning ^31–33^. These findings underscore the need for further study exploring the inflammatory biomarker and metabolic profiles of variable radiologic body composition phenotypes.

Our findings are consistent with emerging observations in critical illness suggesting that tissue-specific body composition metrics provide more granular prognostic information than BMI alone (REFs). BMI as a primary continuous covariate in time to liberation and mortality models showed no association with outcomes. Additionally, BMI-stratified analyses restricted to patients with BMI ≥18.5 kg/m² (n=231) showed no statistically significant differences in rates of successful ventilator liberation or 60 day mortality across normal weight, overweight, and obesity class I and II/III categories. This finding in the context of the significant associations observed for CT-derived metrics reinforces the hypothesis that BMI, by conflating lean and adipose tissue mass into a single measure, obscures tissue-specific contributions to outcomes that volumetric CT quantification can resolve. This is consistent with prior work demonstrating that a higher fat-to-muscle CSA at the carina predicts delayed liberation in IMV patients with hematologic malignancies ^22^. Outside of ARF, metrics similar to MFR have been associated with factors such as insulin resistance and functional capacity supporting their biological plausibility as markers of physiologic reserve ^34,35^. The lack of a BMI-outcome association in ARF is consistent with previous literature where BMI performs inconsistently as a prognostic marker in critically ill patients and supports the clinical value of tissue-specific body composition assessment.

An interesting finding in our study was the J-shaped association between TMM and both delayed liberation from IMV and 60-day mortality, where both extremes of TMM were associated with worse outcomes. The association between low TMM and poor outcomes is consistent with the well-established literature linking sarcopenia and frailty to PMV and critical illness mortality ^36,37^. The association between high TMM and worse outcomes is more counterintuitive, but may be less paradoxical than it appears when interpreted alongside the BMI-stratified data. The BMI-stratified analysis showed that as BMI category increased, SFM rose proportionally more than TMM which resulted in a stepwise decline in MFR across higher BMI strata. This suggests that at the higher end of the TMM distribution, greater absolute muscle mass may simply be a marker of a larger overall body habitus in which adipose accumulation outpaces muscle yielding a less favorable MFR. In this context, it is perhaps the unfavorable MFR rather than high TMM itself that may be driving worse outcomes at the upper extreme which would reinforce that MFR as a more physiologically meaningful discriminator of somatic reserve than either tissue compartment in isolation. Another plausible mechanism that was considered is patients with greater muscle mass may be more prone to patient-ventilator dyssynchrony driven by stronger respiratory muscle effort necessitating deeper sedation or neuromuscular blockade which are associated with ICU-acquired weakness and delayed liberation. In our exploratory NMBA analysis, TMM did not differ significantly between patients who received NMBA and those who did not, though this binary classification did not account for dose or duration of neuromuscular blockade, or for degree of sedation received. This hypothesis deserves prospective evaluation with more detailed sedation data.

The independent association of diabetes with faster ventilator liberation and reduced 60-day mortality across all models warrants cautious interpretation as this finding may reflect confounding by antidiabetic pharmacotherapy rather than a direct protective effect of the disease itself. SGLT2 inhibitors, GLP-1 receptor agonists, and metformin each carry established anti-inflammatory, immunomodulatory, and organ-protective properties that are independent of glucose lowering, with emerging data linking prior SGLT2 inhibitor use to reduced in-hospital mortality in critically ill patients ^38^ and GLP-1 receptor agonists to attenuation of lung injury through decreased cytokine production ^39,40^. Prospective studies with medication usage data will be needed to determine whether the observed benefit is attributable to diabetes, specific antidiabetic agents, or their combination.

Our findings are clinically relevant because thoracic CT imaging is routinely obtained in patients with ARF, creating a practical opportunity to incorporate automated body composition analysis into early risk stratification. CT-derived muscle thoracic body composition metrics could be integrated with quantitative lung parenchymal imaging, physiologic data, and inflammatory biomarkers ^41–43^ and to refine ARF phenotypes and enrich clinical trials testing interventions such as tailored spontaneous breathing trial protocols, early mobilization, or neuromuscular-targeted therapies including diaphragmatic pacing ^12,44,45^. Although our analysis did not include direct measures of diaphragmatic structure or function, established data demonstrate that reduced diaphragm muscle mass and thickness are associated with weaning failure, prolonged ventilation, and higher mortality ^46,47^. This underscores the potential value of incorporating diaphragmatic metrics obtained by ultrasound or CT into future phenotyping frameworks.

Several limitations of this study warrant consideration. First, this is a single-center prospective cohort with a secondary imaging analysis, and generalizability to other institutions and patient populations requires external validation. Second, the modest sample size in this study may generate limited power to detect associations, particularly in subgroup analyses. Third, though we assessed body composition volumetrically from lung apex to base rather than CSA at a single thoracic level, we did not directly evaluate the diaphragm which is the primary muscle of respiration and has been shown to be an important factor in successful weaning. Fourth, we did not assess longitudainal changes in TMM or SFM during the ICU course. These metrics are subject to catabolic stress with significant variation between critically ill patients during the course of their illness. As such, more prognostic information may have been obtained with serial analysis of body composition metrics with CT. Fifth, residual confounding by unmeasured variables including pre-ICU frailty, baseline functional status, nutritional status, and treatment heterogeneity cannot be excluded.

## Conclusions

In this prospective ARF cohort, CT-derived volumetric thoracic body composition metrics, particularly MFR and TMM, are significantly associated with time to ventilator liberation and 60-day mortality. These associations were not captured by BMI stratification, underscoring the value of radiologic phenotyping using tissue-specific, volumetric CT-based assessment. Future prospective, multicenter studies integrating CT-specific body composition metrics with measures of diaphragmatic assessment as well as inflammatory and metabolic biomarkers could yield more granular phenotypes that may benefit from predictive enrichment strategies such as tailored ventilator and rehabilitation protocols to improve outcomes.

## Data Availability

All data produced in the present study are available upon reasonable request to the authors

## Author’s contributions

**Pranav Jain**: Conceptualization, Methodology, Validation, Investigation, Resources, Writing - Original Draft, Writing - Review & Editing, Visualization, Project administration; **Georgios D. Kitsios**: Conceptualization, Methodology, Validation, Formal analysis, Investigation, Resources, Writing - Original Draft, Writing - Review & Editing, Visualization, Supervision, Project administration, Funding acquisition; **Jiantao Pu:** Conceptualization, Methodology, Validation, Investigation, Resources, Writing - Review & Editing, Supervision, Funding acquisition; **Mehdi Nouriae**: Methodology, Validation, Writing - Review & Editing; **Xin Meng, Tong Yu, Jin Wang, Anthony Perti, Grant Kokenberger, Faraaz Shah, William Bain, Bryan J. McVerry, Niall Prendergast**: Investigation, Writing – Review & Editing

## Acknowledgements

The authors thank Ms. Amy Klym for acting as honest broker and obtaining de-identified CT images for analysis. The authors acknowledge the efforts of the ALIR investigators and the bedside clinicians, nurses, and respiratory therapists who assisted in participant enrollment, and thank the patients and their families for their participation in this observational research study.

