## Supplemental Material for "Automated Thoracic Body Composition Analysis on Computed Tomography Is Associated With Ventilator Liberation in Acute Respiratory Failure"

### Contents

#### Supplementary Tables

Supplementary Table 1. Baseline Characteristics of Intubated Patients With and Without CT Scans — p. 3
Supplementary Table 2. Cox Proportional Hazards Models for 60-Day Ventilator Liberation in Patients with Acute Respiratory Failure — p. 4
Supplementary Table 3. Cox Proportional Hazards Models with Restricted Cubic Splines for 60-Day Ventilator Liberation in Patients with Acute Respiratory Failure — p. 6
Supplementary Table 4. Multivariable Logistic Regression Models for 60-Day Mortality in Patients with Acute Respiratory Failure — p. 8
Supplementary Table 5. Logistic Regression Models with Restricted Cubic Splines for 60-Day Mortality in Patients with Acute Respiratory Failure — p. 10
Supplementary Table 6. Multinomial Logistic Regression Models for Ventilator Outcome in Patients with Acute Respiratory Failure — p. 12
Supplementary Table 7. Cox Proportional Hazards Models for 60-Day Ventilator Liberation in ARDS Patients — p. 13
Supplementary Table 8. Cox Proportional Hazards Models with Restricted Cubic Splines for 60-Day Ventilator Liberation in ARDS Patients — p. 14
Supplementary Table 9. Multivariable Logistic Regression Models for 60-Day Mortality in ARDS Patients — p. 15
Supplementary Table 10. Logistic Regression Models with Restricted Cubic Splines for 60-Day Mortality in ARDS Patients — p. 16
Supplementary Table 11. Cox Proportional Hazards and Logistic Regression Models for BMI and 60-Day Ventilator Liberation and Mortality in Patients with Acute Respiratory Failure — p. 17
Supplementary Table 12. Baseline Characteristics by BMI Group — p. 19

#### Supplementary Figures

Supplementary Figure 1. Directed Acyclic Graph (DAG) Depicting the Assumed Causal Relationships Between CT-Derived Body Composition Metrics and IMV Liberation and 60-Day Mortality — p. 20
Supplementary Figure 2. Distribution of Respiratory Failure Etiologies Among Mechanically Ventilated Patients — p. 22
Supplementary Figure 3. Spearman Rank Correlations Between Thoracic Muscle Mass and Respiratory Mechanics at ICU Admission — p. 23
Supplementary Figure 4. Spearman Rank Correlations Between Thoracic Subcutaneous Fat Mass and Respiratory Mechanics at ICU Admission — p. 24
Supplementary Figure 5. Spearman Rank Correlations Between Thoracic Muscle-Fat Ratio and Respiratory Mechanics at ICU Admission — p. 25
Supplementary Figure 6. Spearman Rank Correlations Between Total Lung Volume and Respiratory Mechanics at ICU Admission — p. 26
Supplementary Figure 7. Spearman Rank Correlations Between Consolidated Lung Volume and Respiratory Mechanics at ICU Admission — p. 27

Supplementary Figure 8. Thoracic Muscle Mass Stratified by Neuromuscular Blocking Agent Use During ICU Admission — p. 28
Supplementary Figure 9. Association Between Thoracic Body Composition Metrics and Time to Successful Ventilator Liberation within 60 Days in Patients with Acute Respiratory Distress Syndrome — p. 29
Supplementary Figure 10. Association Between Thoracic Body Composition Metrics and 60-Day Mortality in Patients with Acute Respiratory Distress Syndrome — p. 31
Supplementary Figure 11. Association Between BMI and 60-Day Ventilator Liberation and Mortality: Linear and Restricted Cubic Spline Models — p. 33

**Supplementary Table 1. Baseline Characteristics of Intubated Patients With and Without CT Scans**

| | CT Availability | | |
|---|---|---|---|
| **Characteristic** | **Usable CT** N = 238[1] | **No Usable CT** N = 588[1] | **P-value**[2] |
| **Age (years)** | 59.47 (48.00 - 67.77) | 58.67 (46.66 - 67.95) | 0.712 |
| **Sex** | | | 0.183 |
| Male | 127 (53.4%) | 346 (58.8%) | |
| Female | 111 (46.6%) | 242 (41.2%) | |
| **Body Mass Index (kg/m²)** | 28.65 (24.59 - 34.96) | 30.10 (25.70 - 36.27) | **0.025** |
| **History of Alcohol Use** | 36 (15.1%) | 87 (14.8%) | 0.904 |
| **History of Congestive Heart Failure** | 27 (11.3%) | 82 (13.9%) | 0.317 |
| **History of Chronic Liver Disease** | 14 (5.9%) | 78 (13.3%) | **0.002** |
| **History of COPD** | 54 (22.7%) | 115 (19.6%) | 0.312 |
| **History of Chronic Renal Failure** | 51 (21.4%) | 83 (14.1%) | **0.010** |
| **History of Diabetes** | 76 (31.9%) | 207 (35.2%) | 0.370 |
| **Active Neoplasm** | 14 (5.9%) | 22 (3.7%) | 0.172 |
| **Vasopressors on Intubation** | 155 (65.1%) | 290 (49.3%) | **<0.001** |
| **History of Hypoalbuminemia** | 149 (62.6%) | 266 (45.3%) | **<0.001** |
| Unknown | 0 | 1 | |
| **History of Pulmonary Fibrosis** | 16 (6.7%) | 17 (2.9%) | **0.011** |
| **Hemodynamic Instability on Intubation** | 11 (4.6%) | 18 (3.1%) | 0.270 |
| **History of Immunosuppression** | 55 (23.1%) | 119 (20.2%) | 0.359 |
| **60-Day Mortality** | 83 (34.9%) | 192 (32.7%) | 0.540 |
| **60-Day Successful Liberation** | 181 (76.1%) | 434 (75.5%) | 0.863 |
| Unknown | 0 | 13 | |

[1]Continuous variables presented as median (P25-P75); categorical variables as n (%). P-values from Kruskal-Wallis test for continuous variables and Chi-square test for categorical variables.

[2]Kruskal-Wallis rank sum test; Pearson's Chi-squared test

**Supplementary Table 2. Cox Proportional Hazards Models for 60-Day Ventilator Liberation in Patients with Acute Respiratory Failure**

| Variable | Hazard Ratio (95% CI) | P-value |
|---|---|---|
| **Thoracic Muscle Mass** | | |
| Thoracic Muscle Mass (kg) | 0.96 (0.87–1.05) | 0.357 |
| Age (years) | 0.98 (0.97–0.99) | **<0.001** |
| Sex (Female vs. Male) | 1.09 (0.76–1.56) | 0.656 |
| History of COPD | 0.96 (0.62–1.48) | 0.842 |
| History of Diabetes | 2.03 (1.42–2.88) | **<0.001** |
| History of Chronic Renal Failure | 0.80 (0.51–1.27) | 0.348 |
| History of Immunosuppression | 0.92 (0.62–1.36) | 0.658 |
| History of Hypoalbuminemia | 0.84 (0.61–1.17) | 0.298 |
| **Thoracic Subcutaneous Fat Mass** | | |
| Thoracic Subcutaneous Fat Mass (kg) | 0.95 (0.91–1.00) | 0.067 |
| Age (years) | 0.98 (0.97–0.99) | **0.002** |
| Sex (Female vs. Male) | 1.17 (0.84–1.63) | 0.340 |
| History of COPD | 0.95 (0.61–1.47) | 0.810 |
| History of Diabetes | 2.11 (1.48–3.01) | **<0.001** |
| History of Chronic Renal Failure | 0.77 (0.49–1.22) | 0.270 |
| History of Immunosuppression | 0.90 (0.61–1.33) | 0.596 |
| History of Hypoalbuminemia | 0.83 (0.59–1.15) | 0.251 |
| **Thoracic Muscle-Fat Ratio** | | |
| Thoracic Muscle-Fat Ratio | 1.16 (1.02–1.32) | **0.023** |
| Age (years) | 0.98 (0.97–1.00) | **0.007** |
| Sex (Female vs. Male) | 1.31 (0.93–1.86) | 0.128 |
| History of COPD | 0.93 (0.60–1.44) | 0.730 |
| History of Diabetes | 1.97 (1.38–2.80) | **<0.001** |
| History of Chronic Renal Failure | 0.85 (0.54–1.32) | 0.461 |
| History of Immunosuppression | 0.83 (0.55–1.25) | 0.376 |
| History of Hypoalbuminemia | 0.82 (0.59–1.14) | 0.231 |

*Note: HR > 1 indicates faster liberation (shorter time to extubation); HR < 1 indicates slower liberation. Each model is adjusted for age, sex, history of COPD, history of diabetes, history of*

| Variable | Hazard Ratio (95% CI) | P-value |
|---|---|---|

*chronic renal failure, history of immunosuppression, and history of hypoalbuminemia. Bold p-values indicate statistical significance ($p < 0.05$). CI = Confidence Interval; HR = Hazard Ratio.*

**Supplementary Table 3. Cox Proportional Hazards Models with Restricted Cubic Splines for 60-Day Ventilator Liberation in Patients with Acute Respiratory Failure**

| Variable | Chi-Square | df | P-value |
|---|---|---|---|
| **Thoracic Muscle Mass [N=238, Events=154]** | | | |
| Thoracic Muscle Mass (kg) | 6.05 | 2 | **0.048** |
| Nonlinear Component | 5.50 | 1 | **0.019** |
| Age (years) | 10.61 | 1 | **0.001** |
| Sex (Female vs. Male) | 1.74 | 1 | 0.187 |
| History of COPD | 0.12 | 1 | 0.732 |
| History of Diabetes | 15.85 | 1 | **<0.001** |
| History of Chronic Renal Failure | 0.77 | 1 | 0.382 |
| History of Immunosuppression | 0.21 | 1 | 0.643 |
| History of Hypoalbuminemia | 0.81 | 1 | 0.368 |
| **Thoracic Subcutaneous Fat Mass [N=238, Events=154]** | | | |
| Thoracic Subcutaneous Fat Mass (kg) | 3.42 | 2 | 0.180 |
| Nonlinear Component | 0.02 | 1 | 0.887 |
| Age (years) | 8.97 | 1 | **0.003** |
| Sex (Female vs. Male) | 0.93 | 1 | 0.336 |
| History of COPD | 0.06 | 1 | 0.804 |
| History of Diabetes | 16.86 | 1 | **<0.001** |
| History of Chronic Renal Failure | 1.21 | 1 | 0.271 |
| History of Immunosuppression | 0.30 | 1 | 0.585 |
| History of Hypoalbuminemia | 1.32 | 1 | 0.250 |
| **Thoracic Muscle-Fat Ratio [N=238, Events=154]** | | | |
| Thoracic Muscle-Fat Ratio | 5.93 | 2 | 0.052 |
| Nonlinear Component | 1.04 | 1 | 0.308 |
| Age (years) | 6.45 | 1 | **0.011** |
| Sex (Female vs. Male) | 3.19 | 1 | 0.074 |
| History of COPD | 0.16 | 1 | 0.689 |
| History of Diabetes | 15.13 | 1 | **<0.001** |
| History of Chronic Renal Failure | 0.77 | 1 | 0.380 |

| Variable | Chi-Square | df | P-value |
|---|---|---|---|
| History of Immunosuppression | 0.69 | 1 | 0.407 |
| History of Hypoalbuminemia | 1.44 | 1 | 0.229 |

*Note: Wald chi-square statistics from ANOVA of restricted cubic spline Cox models (3 knots). Each model is adjusted for age, sex, history of COPD, history of diabetes, history of chronic renal failure, history of immunosuppression, and history of hypoalbuminemia. The Nonlinear Component row tests departure from linearity. A significant TOTAL with a non-significant Nonlinear Component supports a linear relationship. Bold p-values indicate statistical significance (p < 0.05). df = degrees of freedom; RCS = restricted cubic splines.*

**Supplementary Table 4. Multivariable Logistic Regression Models for 60-Day Mortality in Patients with Acute Respiratory Failure**

| Variable | Coefficient | Standard Error | P-value | OR (95% CI) |
|---|---|---|---|---|
| **Thoracic Muscle Mass [N=238, Events=83]** | | | | |
| Thoracic Muscle Mass (kg) | 0.110 | 0.083 | 0.185 | 1.12 (0.95–1.31) |
| Age (years) | 0.045 | 0.011 | **<0.001** | 1.05 (1.02–1.07) |
| Sex (Female vs. Male) | -0.148 | 0.330 | 0.653 | 0.86 (0.45–1.65) |
| History of COPD | 0.360 | 0.361 | 0.318 | 1.43 (0.71–2.91) |
| History of Diabetes | -1.314 | 0.363 | **<0.001** | 0.27 (0.13–0.55) |
| History of Chronic Renal Failure | 0.532 | 0.400 | 0.183 | 1.70 (0.78–3.73) |
| History of Immunosuppression | -0.273 | 0.386 | 0.478 | 0.76 (0.36–1.62) |
| History of Hypoalbuminemia | 0.235 | 0.309 | 0.447 | 1.27 (0.69–2.32) |
| **Thoracic Subcutaneous Fat Mass [N=238, Events=83]** | | | | |
| Thoracic Subcutaneous Fat Mass (kg) | 0.084 | 0.043 | **0.049** | 1.09 (1.00–1.18) |
| Age (years) | 0.044 | 0.011 | **<0.001** | 1.05 (1.02–1.07) |
| Sex (Female vs. Male) | -0.333 | 0.309 | 0.281 | 0.72 (0.39–1.31) |
| History of COPD | 0.383 | 0.361 | 0.288 | 1.47 (0.72–2.97) |
| History of Diabetes | -1.407 | 0.369 | **<0.001** | 0.24 (0.12–0.50) |
| History of Chronic Renal Failure | 0.570 | 0.404 | 0.158 | 1.77 (0.80–3.90) |
| History of Immunosuppression | -0.279 | 0.386 | 0.470 | 0.76 (0.35–1.61) |
| History of Hypoalbuminemia | 0.230 | 0.310 | 0.458 | 1.26 (0.69–2.31) |
| **Thoracic Muscle-Fat Ratio [N=238, Events=83]** | | | | |
| Thoracic Muscle-Fat Ratio | -0.416 | 0.226 | 0.066 | 0.66 (0.42–1.03) |
| Age (years) | 0.040 | 0.011 | **<0.001** | 1.04 (1.02–1.06) |
| Sex (Female vs. Male) | -0.476 | 0.320 | 0.137 | 0.62 (0.33–1.16) |
| History of COPD | 0.465 | 0.361 | 0.198 | 1.59 (0.78–3.23) |
| History of Diabetes | -1.396 | 0.367 | **<0.001** | 0.25 (0.12–0.51) |
| History of Chronic Renal Failure | 0.570 | 0.403 | 0.158 | 1.77 (0.80–3.90) |
| History of Immunosuppression | -0.323 | 0.387 | 0.404 | 0.72 (0.34–1.55) |
| History of Hypoalbuminemia | 0.215 | 0.310 | 0.488 | 1.24 (0.68–2.28) |

| Variable | Coefficient | Standard Error | P-value | OR (95% CI) |
|---|---|---|---|---|

*Note: Each model is adjusted for age, sex, history of COPD, history of diabetes, history of chronic renal failure, history of immunosuppression, and history of hypoalbuminemia. OR > 1 indicates increased odds of 60-day mortality; OR < 1 indicates decreased odds. Confidence intervals calculated using ±1.96×SE (Wald method). Bold p-values indicate statistical significance ($p < 0.05$). CI = Confidence Interval; OR = Odds Ratio; SE = Standard Error.*

**Supplementary Table 5. Logistic Regression Models with Restricted Cubic Splines for 60-Day Mortality in Patients with Acute Respiratory Failure**

| Variable | Chi-Square | df | P-value |
|---|---|---|---|
| **Thoracic Muscle Mass [N=238, Events=83]** | | | |
| Thoracic Muscle Mass (kg) | 6.81 | 2 | **0.033** |
| Nonlinear Component | 5.36 | 1 | **0.021** |
| Age (years) | 16.63 | 1 | **<0.001** |
| Sex (Female vs. Male) | 1.60 | 1 | 0.206 |
| History of COPD | 1.44 | 1 | 0.230 |
| History of Diabetes | 13.76 | 1 | **<0.001** |
| History of Chronic Renal Failure | 1.82 | 1 | 0.177 |
| History of Immunosuppression | 0.56 | 1 | 0.453 |
| History of Hypoalbuminemia | 0.43 | 1 | 0.511 |
| **Thoracic Subcutaneous Fat Mass [N=238, Events=83]** | | | |
| Thoracic Subcutaneous Fat Mass (kg) | 3.86 | 2 | 0.145 |
| Nonlinear Component | 0.00 | 1 | 0.978 |
| Age (years) | 16.26 | 1 | **<0.001** |
| Sex (Female vs. Male) | 1.16 | 1 | 0.282 |
| History of COPD | 1.12 | 1 | 0.289 |
| History of Diabetes | 14.53 | 1 | **<0.001** |
| History of Chronic Renal Failure | 1.99 | 1 | 0.158 |
| History of Immunosuppression | 0.52 | 1 | 0.469 |
| History of Hypoalbuminemia | 0.55 | 1 | 0.458 |
| **Thoracic Muscle-Fat Ratio [N=238, Events = 83]** | | | |
| Thoracic Muscle-Fat Ratio | 4.20 | 2 | 0.123 |
| Nonlinear Component | 0.42 | 1 | 0.517 |
| Age (years) | 13.26 | 1 | **<0.001** |
| Sex (Female vs. Male) | 2.60 | 1 | 0.107 |
| History of COPD | 1.68 | 1 | 0.195 |
| History of Diabetes | 14.79 | 1 | **<0.001** |
| History of Chronic Renal Failure | 2.00 | 1 | 0.157 |

| Variable | Chi-Square | df | P-value |
| --- | --- | --- | --- |
| History of Immunosuppression | 0.64 | 1 | 0.422 |
| History of Hypoalbuminemia | 0.46 | 1 | 0.499 |

*Note: Wald chi-square statistics from ANOVA of restricted cubic spline logistic regression models (3 knots, placed at 10th, 50th, and 90th percentiles). Each model is adjusted for age, sex, history of COPD, history of diabetes, history of chronic renal failure, history of immunosuppression, and history of hypoalbuminemia. The Nonlinear Component row tests departure from linearity. A significant TOTAL with a non-significant Nonlinear Component supports a linear relationship on the log-odds scale. Bold p-values indicate statistical significance (p < 0.05). df = degrees of freedom; RCS = restricted cubic splines.*

**Supplementary Table 6. Multinomial Logistic Regression Models for Ventilator Outcome in Patients with Acute Respiratory Failure**

| Outcome | Variable | OR (95% CI) | P-value |
|---|---|---|---|
| **Thoracic Muscle Mass** | | | |
| PMV | Thoracic Muscle Mass (kg) | 0.72 (0.51–1.03) | 0.074 |
| PMV | Age (years) | 0.97 (0.95–1.00) | **0.025** |
| PMV | Sex (Female vs. Male) | 0.64 (0.25–1.65) | 0.357 |
| Non-Survivor | Thoracic Muscle Mass (kg) | 1.09 (0.93–1.28) | 0.299 |
| Non-Survivor | Age (years) | 1.04 (1.02–1.06) | **<0.001** |
| Non-Survivor | Sex (Female vs. Male) | 0.90 (0.49–1.67) | 0.742 |
| **Thoracic Subcutaneous Fat Mass** | | | |
| PMV | Thoracic Subcutaneous Fat Mass (kg) | 0.95 (0.82–1.10) | 0.476 |
| PMV | Age (years) | 0.98 (0.95–1.00) | 0.060 |
| PMV | Sex (Female vs. Male) | 1.00 (0.44–2.30) | 0.996 |
| Non-Survivor | Thoracic Subcutaneous Fat Mass (kg) | 1.05 (0.97–1.14) | 0.203 |
| Non-Survivor | Age (years) | 1.04 (1.02–1.06) | **<0.001** |
| Non-Survivor | Sex (Female vs. Male) | 0.79 (0.44–1.40) | 0.420 |
| **Thoracic Muscle-Fat Ratio** | | | |
| PMV | Thoracic Muscle-Fat Ratio | 0.71 (0.43–1.18) | 0.189 |
| PMV | Age (years) | 0.97 (0.94–0.99) | **0.018** |
| PMV | Sex (Female vs. Male) | 0.81 (0.34–1.94) | 0.643 |
| Non-Survivor | Thoracic Muscle-Fat Ratio | 0.72 (0.48–1.07) | 0.102 |
| Non-Survivor | Age (years) | 1.03 (1.01–1.05) | **0.003** |
| Non-Survivor | Sex (Female vs. Male) | 0.69 (0.38–1.26) | 0.227 |

*Note: Reference category = Early Liberation. Each model is adjusted for age and sex. OR > 1 indicates increased odds of the outcome category relative to early liberation; OR < 1 indicates decreased odds. Confidence intervals calculated using ±1.96×SE (Wald method). Bold p-values indicate statistical significance (p < 0.05). PMV = Prolonged Mechanical Venitlation; CI = Confidence Interval; OR = Odds Ratio; SE = Standard Error.*

**Supplementary Table 7. Cox Proportional Hazards Models for 60-Day Ventilator Liberation in ARDS Patients**

| Variable | Hazard Ratio (95% CI) | P-value |
|---|---|---|
| **Model 1: Thoracic Muscle Mass [N=81, Events=57]** | | |
| Thoracic Muscle Mass (kg) | 0.85 (0.72–1.02) | 0.077 |
| Age (years) | 0.98 (0.96–0.99) | **0.005** |
| **Model 2: Thoracic Subcutaneous Fat Mass [N=81, Events=57]** | | |
| Thoracic Subcutaneous Fat Mass (kg) | 0.92 (0.83–1.03) | 0.141 |
| Age (years) | 0.98 (0.97–1.00) | **0.023** |
| **Model 3: Thoracic Muscle-Fat Ratio [N=81, Events=57]** | | |
| Thoracic Muscle-Fat Ratio | 1.05 (0.85–1.30) | 0.629 |
| Age (years) | 0.98 (0.97–1.00) | **0.014** |

*Note: HR > 1 indicates faster liberation (shorter time to extubation); HR < 1 indicates slower liberation. Each model is adjusted for age only. Bold p-values indicate statistical significance (p < 0.05). CI = Confidence Interval; HR = Hazard Ratio.*

**Supplementary Table 8. Cox Proportional Hazards Models with Restricted Cubic Splines for 60-Day Ventilator Liberation in ARDS Patients**

| Variable | Chi-Square | df | P-value |
|---|---|---|---|
| **Model 1: Thoracic Muscle Mass [N=81, Events=57]** | | | |
| Thoracic Muscle Mass (kg) | 4.40 | 2 | 0.111 |
| Nonlinear Component | 2.81 | 1 | 0.094 |
| Age (years) | 8.87 | 1 | **0.003** |
| TOTAL | 12.10 | 3 | **0.007** |
| **Model 2: Thoracic Subcutaneous Fat Mass [N=81, Events=57]** | | | |
| Thoracic Subcutaneous Fat Mass (kg) | 2.76 | 2 | 0.252 |
| Nonlinear Component | 1.20 | 1 | 0.273 |
| Age (years) | 5.67 | 1 | **0.017** |
| TOTAL | 9.38 | 3 | **0.025** |
| **Model 3: Thoracic Muscle-Fat Ratio [N=81, Events=57]** | | | |
| Thoracic Muscle-Fat Ratio | 0.28 | 2 | 0.870 |
| Nonlinear Component | 0.03 | 1 | 0.855 |
| Age (years) | 5.80 | 1 | **0.016** |
| TOTAL | 7.15 | 3 | 0.067 |

*Note: Wald chi-square statistics from ANOVA of restricted cubic spline Cox models (3 knots). Each model is adjusted for age only. The Nonlinear Component row tests departure from linearity. A significant TOTAL with a non-significant Nonlinear Component supports a linear relationship. Bold p-values indicate statistical significance (p < 0.05). df = degrees of freedom; RCS = restricted cubic splines.*

**Supplementary Table 9. Multivariable Logistic Regression Models for 60-Day Mortality in ARDS Patients**

| Variable | Coefficient | Standard Error | P-value | OR (95% CI) |
|---|---|---|---|---|
| **Model 1: Thoracic Muscle Mass [N=81, Events=23]** | | | | |
| Thoracic Muscle Mass (kg) | 0.374 | 0.178 | **0.036** | 1.45 (1.03–2.06) |
| Age (years) | 0.056 | 0.019 | **0.004** | 1.06 (1.02–1.10) |
| **Model 2: Thoracic Subcutaneous Fat Mass [N=81, Events=23]** | | | | |
| Thoracic Subcutaneous Fat Mass (kg) | 0.174 | 0.101 | 0.085 | 1.19 (0.98–1.45) |
| Age (years) | 0.046 | 0.019 | **0.013** | 1.05 (1.01–1.09) |
| **Model 3: Thoracic Muscle-Fat Ratio [N=81, Events=23]** | | | | |
| Thoracic Muscle-Fat Ratio | -0.383 | 0.476 | 0.420 | 0.68 (0.27–1.73) |
| Age (years) | 0.044 | 0.018 | **0.017** | 1.04 (1.01–1.08) |

*Note: Each model is adjusted for age only. OR > 1 indicates increased odds of 60-day mortality; OR < 1 indicates decreased odds. Confidence intervals calculated using ±1.96×SE (Wald method). Bold p-values indicate statistical significance ($p < 0.05$). CI = Confidence Interval; OR = Odds Ratio; SE = Standard Error.*

**Supplementary Table 10. Logistic Regression Models with Restricted Cubic Splines for 60-Day Mortality in ARDS Patients**

| Variable | Chi-Square | df | P-value |
|---|---|---|---|
| **Model 1: Thoracic Muscle Mass [N=81, Events=23]** | | | |
| Thoracic Muscle Mass (kg) | 6.85 | 2 | **0.032** |
| Nonlinear Component | 4.37 | 1 | **0.037** |
| Age (years) | 9.05 | 1 | **0.003** |
| TOTAL | 11.59 | 3 | **0.009** |
| **Model 2: Thoracic Subcutaneous Fat Mass [N=81, Events=23]** | | | |
| Thoracic Subcutaneous Fat Mass (kg) | 3.87 | 2 | 0.145 |
| Nonlinear Component | 1.55 | 1 | 0.213 |
| Age (years) | 6.80 | 1 | **0.009** |
| TOTAL | 9.58 | 3 | **0.022** |
| **Model 3: Thoracic Muscle-Fat Ratio [N=81, Events=23]** | | | |
| Thoracic Muscle-Fat Ratio | 1.16 | 2 | 0.561 |
| Nonlinear Component | 1.02 | 1 | 0.312 |
| Age (years) | 5.83 | 1 | **0.016** |
| TOTAL | 6.95 | 3 | 0.074 |

*Note: Wald chi-square statistics from ANOVA of restricted cubic spline logistic regression models (3 knots, placed at 10th, 50th, and 90th percentiles). Each model is adjusted for age only. The Nonlinear Component row tests departure from linearity. A significant TOTAL with a non-significant Nonlinear Component supports a linear relationship on the log-odds scale. Bold p-values indicate statistical significance ($p < 0.05$). df = degrees of freedom; RCS = restricted cubic splines.*

**Supplementary Table 11. Cox Proportional Hazards and Logistic Regression Models for BMI and 60-Day Ventilator Liberation and Mortality in Patients with Acute Respiratory Failure**

| Variable | Effect (95% CI) | P-value |
|---|---|---|
| **Model 1: Cox Proportional Hazards – Linear BMI (60-Day Liberation)** | | |
| BMI (kg/m²) | 1.00 (0.98–1.02) | 0.803 |
| Age (years) | 0.98 (0.97–0.99) | **<0.001** |
| Sex (Female vs. Male) | 1.16 (0.84–1.62) | 0.369 |
| History of COPD | 0.94 (0.60–1.46) | 0.780 |
| History of Chronic Renal Failure | 0.82 (0.52–1.29) | 0.384 |
| History of Diabetes | 2.02 (1.42–2.88) | **<0.001** |
| History of Immunosuppression | 0.92 (0.62–1.37) | 0.673 |
| History of Hypoalbuminemia | 0.84 (0.60–1.16) | 0.293 |
| **Model 2: Cox Proportional Hazards – Restricted Cubic Spline BMI (60-Day Liberation)** | | |
| BMI (kg/m²) | Overall association P = 0.923 | Nonlinear P = 0.749 |
| Age (years) | 0.98 (0.97–0.99) | **<0.001** |
| Sex (Female vs. Male) | 1.16 (0.84–1.62) | 0.369 |
| History of COPD | 0.94 (0.60–1.46) | 0.780 |
| History of Chronic Renal Failure | 0.82 (0.52–1.29) | 0.384 |
| History of Diabetes | 2.02 (1.42–2.88) | **<0.001** |
| History of Immunosuppression | 0.92 (0.62–1.37) | 0.673 |
| History of Hypoalbuminemia | 0.84 (0.60–1.16) | 0.293 |
| **Model 3: Logistic Regression – Linear BMI (60-Day Mortality)** | | |
| BMI (kg/m²) | 1.01 (0.98–1.04) | 0.589 |
| Age (years) | 1.05 (1.02–1.07) | **<0.001** |
| Sex (Female vs. Male) | 0.74 (0.40–1.34) | 0.315 |
| History of COPD | 1.52 (0.75–3.08) | 0.243 |
| History of Chronic Renal Failure | 1.70 (0.77–3.74) | 0.186 |
| History of Diabetes | 0.27 (0.13–0.53) | **<0.001** |
| History of Immunosuppression | 0.75 (0.35–1.58) | 0.457 |
| History of Hypoalbuminemia | 1.26 (0.69–2.33) | 0.448 |

| Variable | Effect (95% CI) | P-value |
|---|---|---|
| **Model 4: Logistic Regression – Restricted Cubic Spline BMI (60-Day Mortality)** | | |
| BMI (kg/m²) | Overall association P = 0.821 | Nonlinear P = 0.747 |
| Age (years) | 1.05 (1.02–1.07) | **<0.001** |
| Sex (Female vs. Male) | 0.74 (0.40–1.34) | 0.315 |
| History of COPD | 1.52 (0.75–3.08) | 0.243 |
| History of Chronic Renal Failure | 1.70 (0.77–3.74) | 0.186 |
| History of Diabetes | 0.27 (0.13–0.53) | **<0.001** |
| History of Immunosuppression | 0.75 (0.35–1.58) | 0.457 |
| History of Hypoalbuminemia | 1.26 (0.69–2.33) | 0.448 |

*Note: For linear models, HR/OR > 1 indicates faster liberation or increased mortality risk, respectively; HR/OR < 1 indicates the opposite. For restricted cubic spline (RCS) models, the BMI row reports the overall association p-value (test of BMI's total contribution to the model) and the nonlinearity p-value (test of departure from linearity) rather than a single effect estimate, since spline terms do not correspond to a single interpretable HR/OR. All models are adjusted for age, sex, history of COPD, history of diabetes, history of chronic renal failure, history of immunosuppression, and history of hypoalbuminemia. Bold p-values indicate statistical significance (p < 0.05). CI = Confidence Interval; HR = Hazard Ratio; OR = Odds Ratio.*

**Supplementary Table 12. Baseline Characteristics by BMI Group**

| | BMI Group | | | | |
|---|---|---|---|---|---|
| **Characteristic** | **Normal** N = 56[1] | **Overweight** N = 70[1] | **Obesity Class 1** N = 46[1] | **Obesity Class 2/3** N = 59[1] | **p-value**[2] |
| **Age (years)** | 53.33 (18.96) | 61.48 (15.89) | 58.31 (15.64) | 55.83 (13.90) | **0.036** |
| **Sex** | | | | | 0.916 |
| Male | 32 (57.1%) | 38 (54.3%) | 24 (52.2%) | 30 (50.8%) | |
| Female | 24 (42.9%) | 32 (45.7%) | 22 (47.8%) | 29 (49.2%) | |
| **Thoracic Muscle Mass (kg)** | 3.46 (1.24) | 3.78 (1.66) | 4.52 (2.15) | 4.56 (2.18) | **0.002** |
| **Subcutaneous Fat Mass (kg)** | 2.41 (1.57) | 3.98 (2.59) | 5.86 (3.30) | 7.41 (3.69) | **<0.001** |
| **Muscle-to-Fat Ratio** | 1.96 (1.66) | 1.13 (0.57) | 0.89 (0.40) | 0.66 (0.23) | **<0.001** |
| **History of Alcohol Use** | 8 (14.3%) | 12 (17.1%) | 5 (10.9%) | 10 (16.9%) | 0.788 |
| **History of Congestive Heart Failure** | 7 (12.5%) | 8 (11.4%) | 6 (13.0%) | 5 (8.5%) | 0.875 |
| **History of Chronic Liver Disease** | 3 (5.4%) | 4 (5.7%) | 2 (4.3%) | 5 (8.5%) | 0.824 |
| **History of COPD** | 14 (25.0%) | 16 (22.9%) | 12 (26.1%) | 10 (16.9%) | 0.662 |
| **History of Chronic Renal Failure** | 16 (28.6%) | 18 (25.7%) | 11 (23.9%) | 6 (10.2%) | 0.076 |
| **History of Diabetes** | 10 (17.9%) | 27 (38.6%) | 15 (32.6%) | 22 (37.3%) | 0.063 |
| **Active Neoplasm** | 4 (7.1%) | 3 (4.3%) | 0 (0.0%) | 7 (11.9%) | 0.073 |
| **History of Hypoalbuminemia** | 36 (64.3%) | 42 (60.0%) | 26 (56.5%) | 40 (67.8%) | 0.645 |
| **History of Pulmonary Fibrosis** | 7 (12.5%) | 6 (8.6%) | 2 (4.3%) | 1 (1.7%) | 0.113 |
| **History of Immunosuppression** | 19 (33.9%) | 17 (24.3%) | 10 (21.7%) | 8 (13.6%) | 0.080 |
| **Hemodynamic Instability on Intubation** | 3 (5.4%) | 2 (2.9%) | 5 (10.9%) | 1 (1.7%) | 0.132 |
| **CRRT on Intubation** | 0 (0.0%) | 2 (2.9%) | 1 (2.2%) | 1 (1.7%) | 0.668 |
| **Vasopressors on Intubation** | 30 (53.6%) | 49 (70.0%) | 32 (69.6%) | 38 (64.4%) | 0.225 |
| **Worst P/F Ratio (24hr)** | 155.75 (112.43, 206.50) | 164.00 (116.67, 205.00) | 174.00 (117.14, 212.50) | 155.00 (105.00, 234.29) | 0.933 |
| **60-Day Successful Liberation** | 39 (69.6%) | 39 (55.7%) | 31 (67.4%) | 40 (67.8%) | 0.327 |
| **60-Day Mortality** | 17 (30.4%) | 30 (42.9%) | 15 (32.6%) | 19 (32.2%) | 0.432 |

[1]Mean (SD); n (%); Median (Q1, Q3)

[2]One-way analysis of means; Pearson's Chi-squared test; Kruskal-Wallis rank sum test

**Supplementary Figure 1. Directed Acyclic Graph (DAG) Depicting the Assumed Causal Relationships Between CT-Derived Body Composition Metrics and IMV Liberation and 60-Day Mortality.**

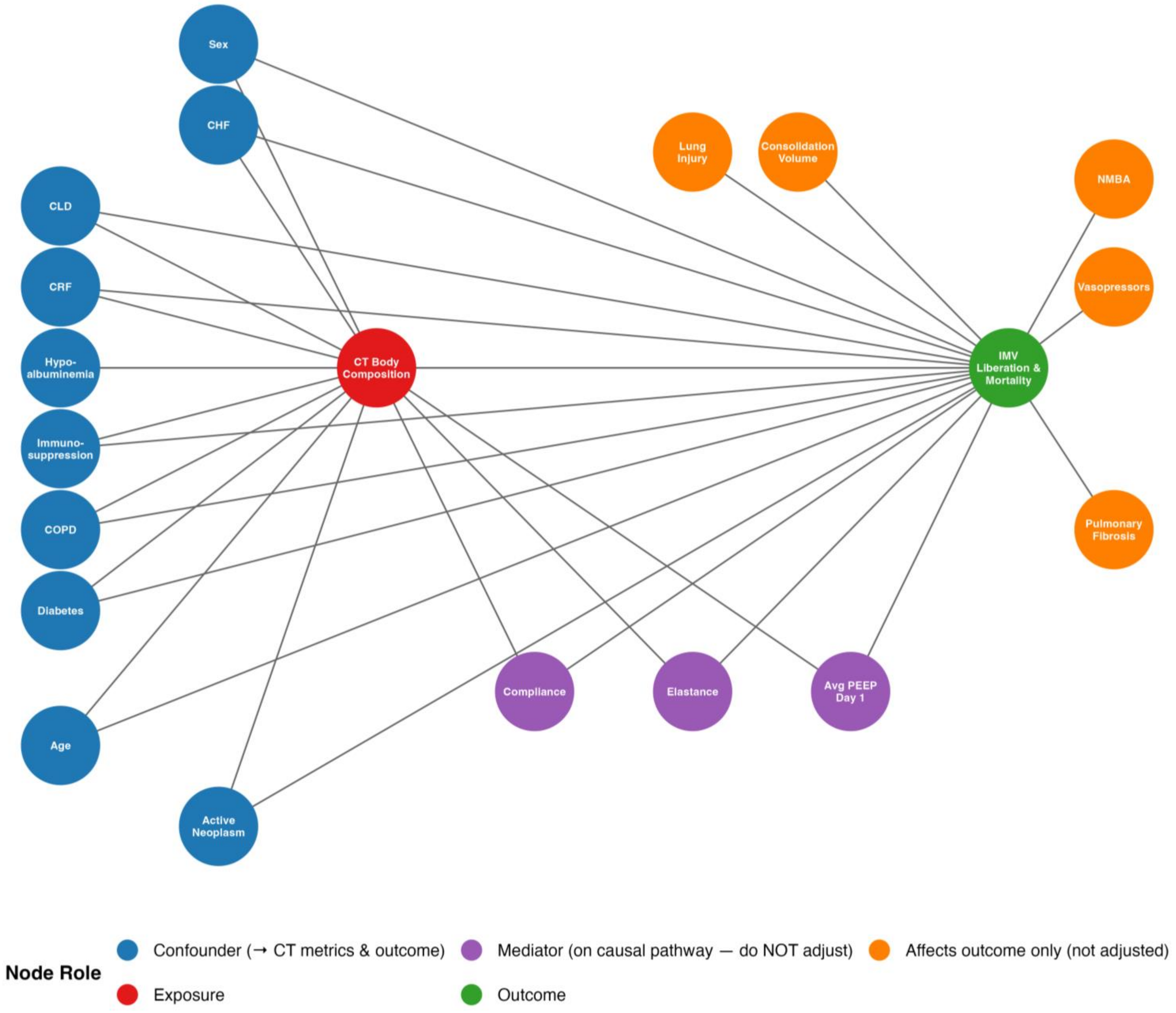


Nodes represent variables included in the causal model. CT Body Composition (red) is the exposure of interest and IMV Liberation & Mortality (green) is the primary outcome. Confounders (blue) are variables with paths to both the exposure and outcome and were included as covariates in multivariable models. CHF, CLD, and active neoplasm were not adjusted for given low proportion of patients with these comorbidities and to maintain events per variable > 10. Outcome-only variables (orange) direct paths to the outcome alone and were not adjusted for. Mediators (purple) lie on the causal pathway and were excluded from adjusted models. Arrows represent assumed directed causal effects

Abbreviations: CRF, chronic renal failure; CHF, congestive heart failure; CLD, chronic liver disease; COPD, chronic obstructive pulmonary disease; CT, computed tomography; DAG, directed acyclic graph; IMV, invasive mechanical ventilation; NMBA, neuromuscular blocking agent; PEEP, positive end-expiratory pressure.

**Supplementary Figure 2. Distribution of Respiratory Failure Etiologies Among Mechanically Ventilated Patients.**

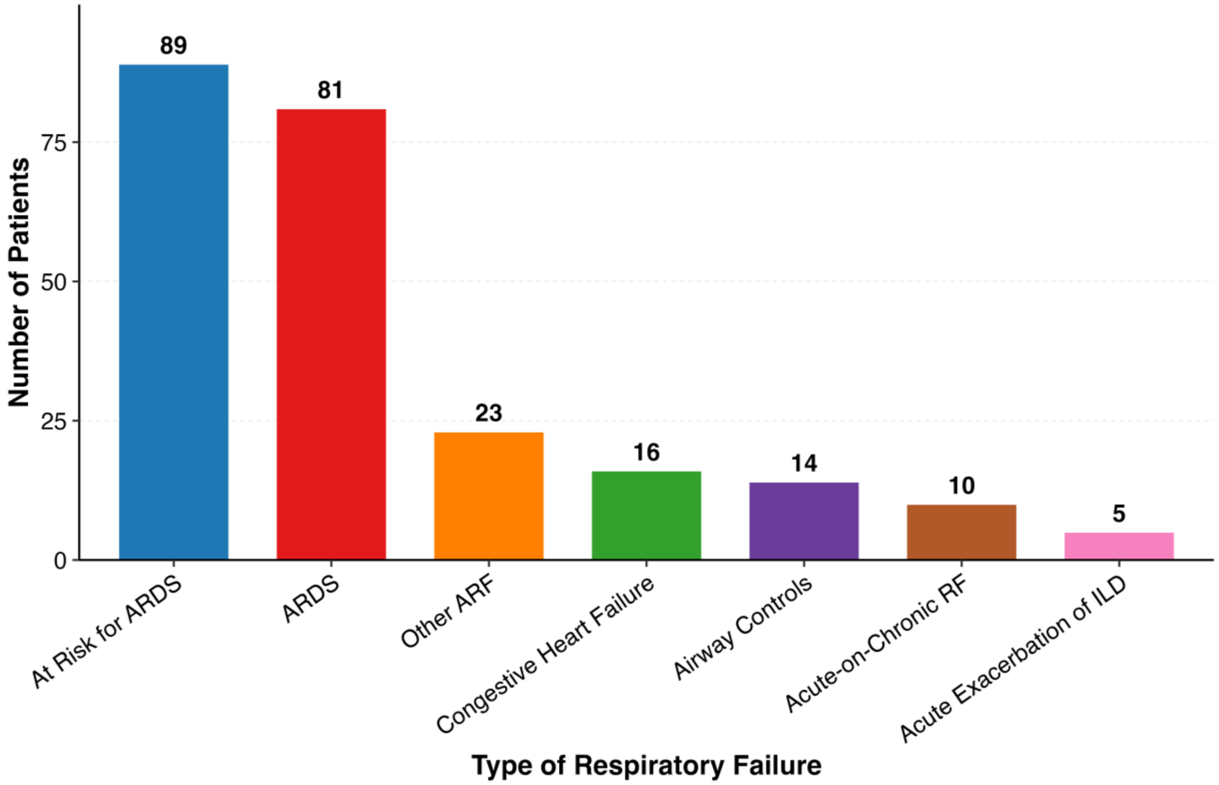


Bar chart depicting the frequency of each respiratory failure etiology among patients included in the study cohort. Each bar represents one diagnostic category, ordered by decreasing frequency. The number of patients in each category is displayed above the corresponding bar.

Abbreviations: ARDS, acute respiratory distress syndrome; ARF, acute respiratory failure; CHF, congestive heart failure; ILD, interstitial lung disease; RF, respiratory failure.

**Supplementary Figure 3. Spearman Rank Correlations Between Thoracic Muscle Mass and Respiratory Mechanics at ICU Admission.**

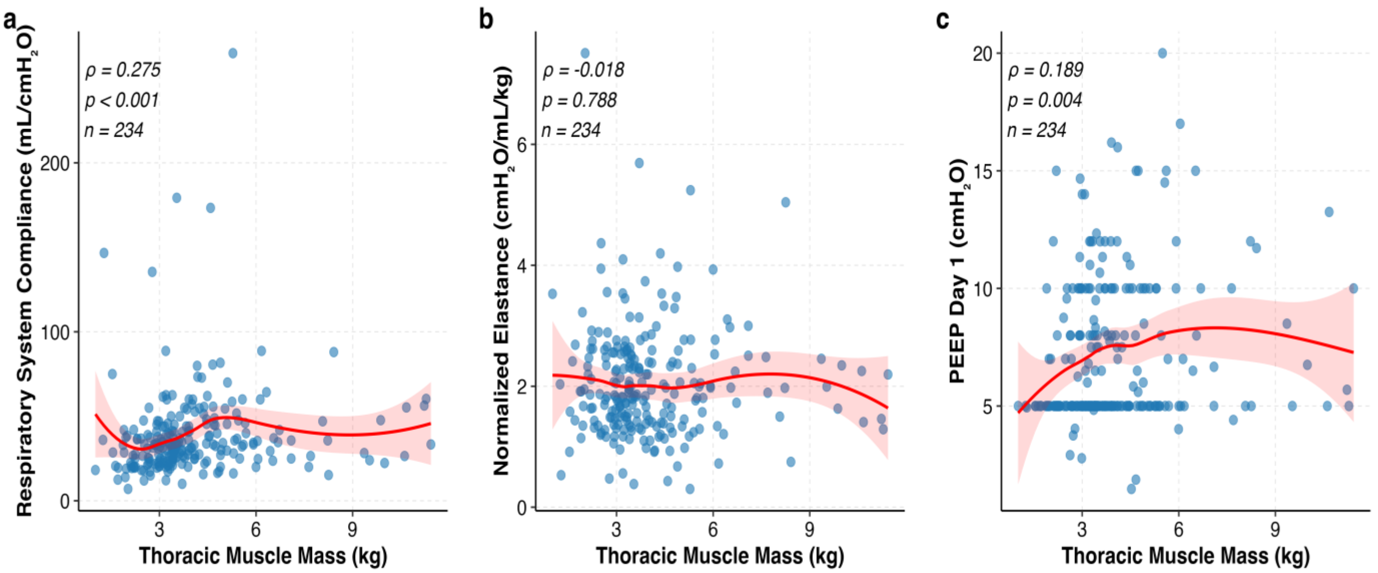


Scatter plots depicting the associations between CT-derived thoracic muscle mass (kg) and (a) respiratory system compliance (mL/cm$H_2O$), (b) normalized respiratory system elastance (cm$H_2O$/mL/kg), and (c) mean positive end-expiratory pressure (PEEP) on Day 1 of mechanical ventilation (cm$H_2O$). Each point represents an individual patient. The red line represents a locally estimated scatterplot smoothing (LOESS) curve with 95% confidence interval (shaded region) to illustrate the non-parametric trend. Spearman's rank correlation coefficient (ρ), corresponding p-value, and sample size (n) are displayed in the upper left corner of each panel. CT, computed tomography; ICU, intensive care unit; PEEP, positive end-expiratory pressure.

**Supplementary Figure 4. Spearman Rank Correlations Between Thoracic Subcutaneous Fat Mass and Respiratory Mechanics at ICU Admission.**

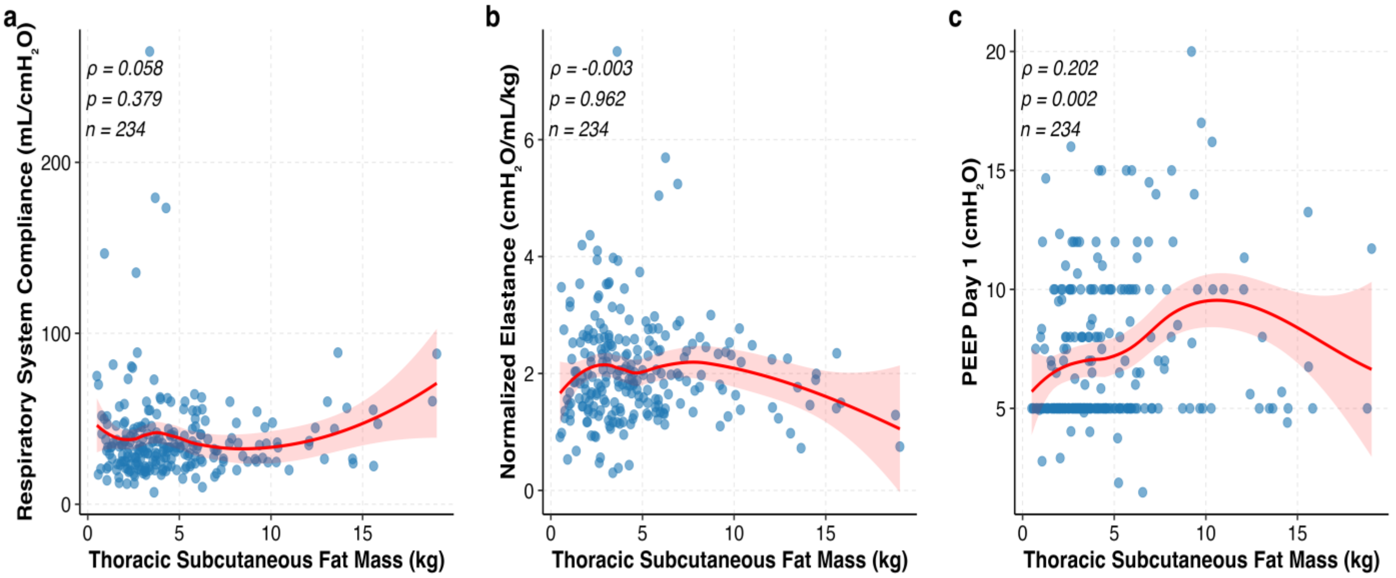


Scatter plots depicting the associations between CT-derived thoracic subcutaneous fat mass (kg) and (a) respiratory system compliance (mL/cm$H_2O$), (b) normalized respiratory system elastance (cm$H_2O$/mL/kg), and (c) mean positive end-expiratory pressure (PEEP) on Day 1 of mechanical ventilation (cm$H_2O$). Each point represents an individual patient. The red line represents a locally estimated scatterplot smoothing (LOESS) curve with 95% confidence interval (shaded region) to illustrate the non-parametric trend. Spearman's rank correlation coefficient (ρ), corresponding p-value, and sample size (n) are displayed in the upper left corner of each panel. CT, computed tomography; ICU, intensive care unit; PEEP, positive end-expiratory pressure.

**Supplementary Figure 5. Spearman Rank Correlations Between Thoracic Muscle-Fat Ratio and Respiratory Mechanics at ICU Admission.**

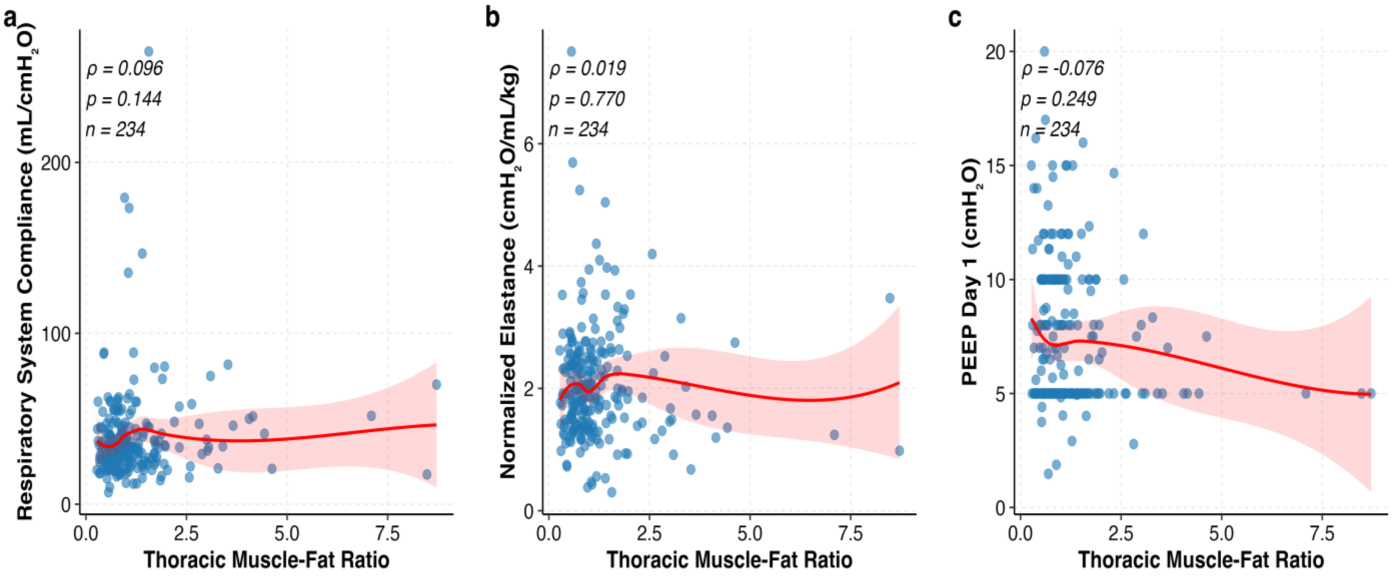


Scatter plots depicting the associations between CT-derived thoracic muscle-fat ratio and (a) respiratory system compliance (mL/cm$H_2O$), (b) normalized respiratory system elastance (cm$H_2O$/mL/kg), and (c) mean positive end-expiratory pressure (PEEP) on Day 1 of mechanical ventilation (cm$H_2O$). Each point represents an individual patient. The red line represents a locally estimated scatterplot smoothing (LOESS) curve with 95% confidence interval (shaded region) to illustrate the non-parametric trend. Spearman's rank correlation coefficient (ρ), corresponding p-value, and sample size (n) are displayed in the upper left corner of each panel. CT, computed tomography; ICU, intensive care unit; PEEP, positive end-expiratory pressure.

**Supplementary Figure 6. Spearman Rank Correlations Between Total Lung Volume and Respiratory Mechanics at ICU Admission.**

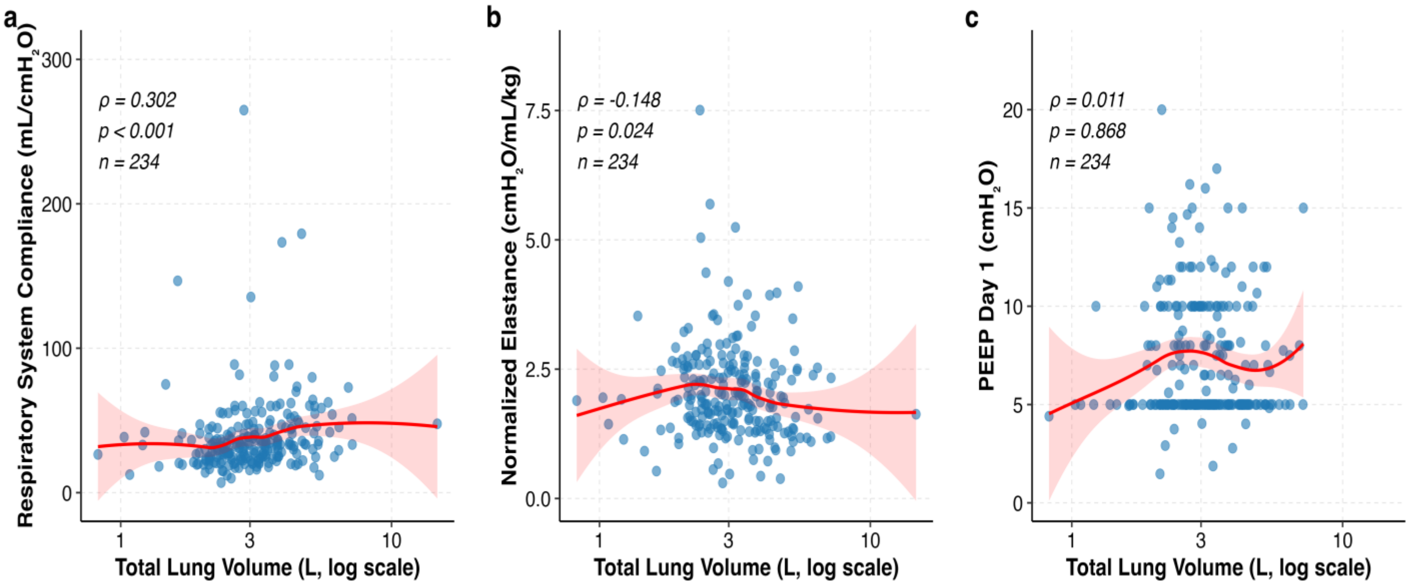


Scatter plots depicting the associations between CT-derived total lung volume (L) and (a) respiratory system compliance (mL/cm$H_2O$), (b) normalized respiratory system elastance (cm$H_2O$/mL/kg), and (c) mean positive end-expiratory pressure (PEEP) on Day 1 of mechanical ventilation (cm$H_2O$). X-axis log transformed for skewed data. Each point represents an individual patient. The red line represents a locally estimated scatterplot smoothing (LOESS) curve with 95% confidence interval (shaded region) to illustrate the non-parametric trend. Spearman's rank correlation coefficient (ρ), corresponding p-value, and sample size (n) are displayed in the upper left corner of each panel. CT, computed tomography; ICU, intensive care unit; PEEP, positive end-expiratory pressure.

**Supplementary Figure 7. Spearman Rank Correlations Between Consolidated Lung Volume and Respiratory Mechanics at ICU Admission.**

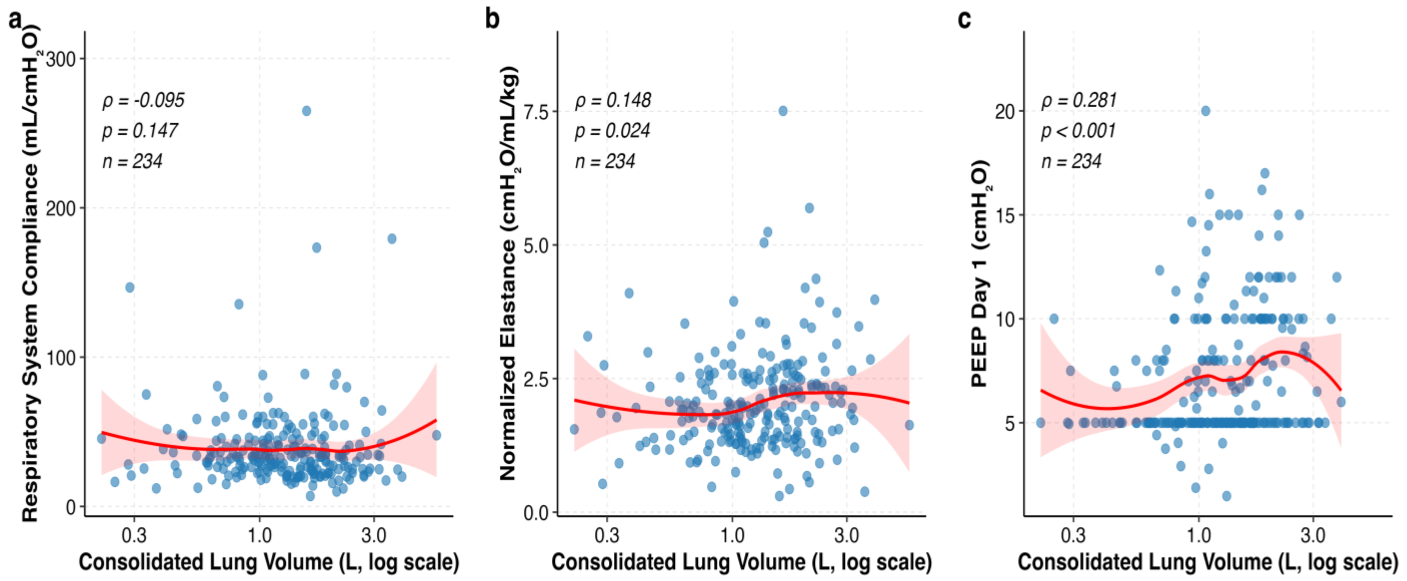


Scatter plots depicting the associations between CT-derived consolidated lung volume (L) and (a) respiratory system compliance (mL/cmH$_2$O), (b) normalized respiratory system elastance (cmH$_2$O/mL/kg), and (c) mean positive end-expiratory pressure (PEEP) on Day 1 of mechanical ventilation (cmH$_2$O). X-axis log transformed for skewed data. Each point represents an individual patient. The red line represents a locally estimated scatterplot smoothing (LOESS) curve with 95% confidence interval (shaded region) to illustrate the non-parametric trend. Spearman's rank correlation coefficient (ρ), corresponding p-value, and sample size (n) are displayed in the upper left corner of each panel. CT, computed tomography; ICU, intensive care unit; PEEP, positive end-expiratory pressure.

**Supplementary Figure 8. Thoracic Muscle Mass Stratified by Neuromuscular Blocking Agent Use During ICU Admission.**

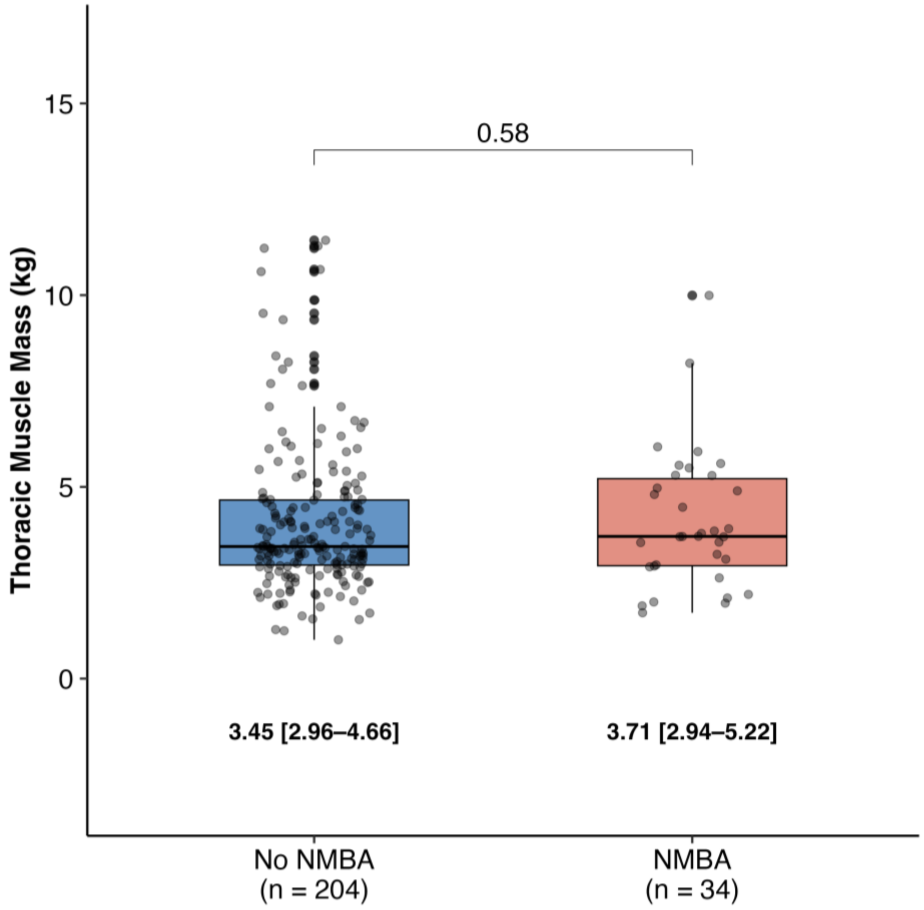


Box and whisker plots comparing CT-derived thoracic muscle mass (kg) between patients who received neuromuscular blocking agents (NMBA) during their ICU stay and those who did not. Individual patient values are overlaid as jittered points. The horizontal line within each box represents the median, box boundaries represent the interquartile range (IQR), and whiskers extend to 1.5 times the IQR. Median [Q1–Q3] values are displayed below each group. Group sample sizes are indicated on the x-axis. Between-group comparison was performed using the Wilcoxon rank-sum test with the p-value displayed above the comparison bracket. CT, computed tomography; ICU, intensive care unit; IQR, interquartile range; NMBA, neuromuscular blocking agent; Q1, first quartile; Q3, third quartile.

**Supplementary Figure 9. Association Between Thoracic Body Composition Metrics and Time to Successful Ventilator Liberation within 60 Days in Patients with Acute Respiratory Distress Syndrome.**

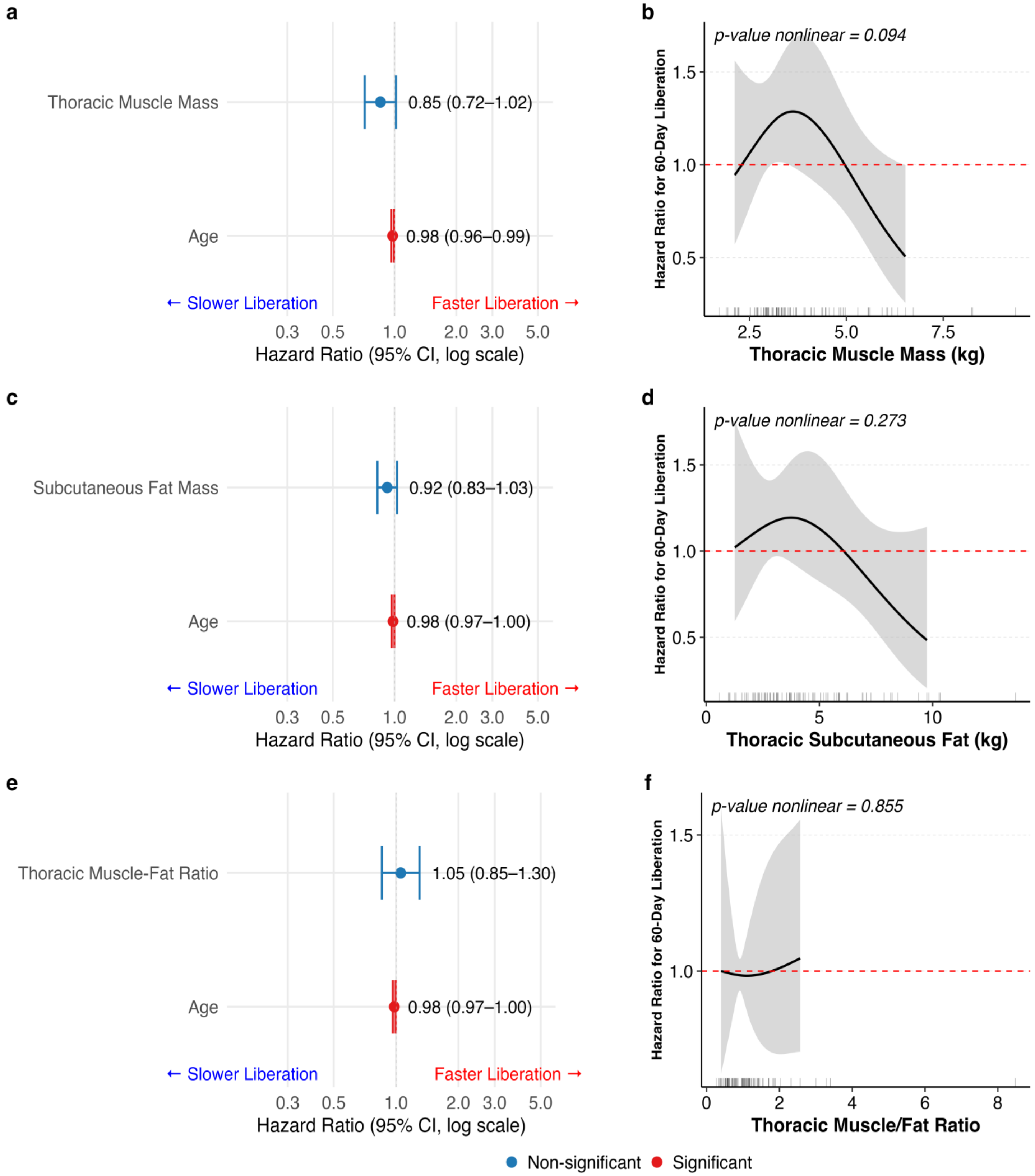

Panels (a), (c), and (e) display forest plots from multivariable Cox proportional hazards regression models examining the association of thoracic muscle mass (a), thoracic subcutaneous fat mass (c), and thoracic muscle-to-fat ratio (e) with 60-day ventilator liberation, adjusted for age in ARDS. Each point represents the hazard ratio (HR) with 95% confidence intervals (CI); values greater than 1 indicate faster liberation and values less than 1 indicate slower liberation. Red points and error bars denote statistically significant associations ($p < 0.05$); blue denotes non-significant associations. Panels (b), (d), and (f) display restricted cubic spline (RCS) curves from corresponding Cox regression models, illustrating the dose-response relationship between each body composition metric and the hazard of liberation. The shaded region represents the 95% CI. The dashed red horizontal line indicates a hazard ratio of 1 (no effect). Rug marks along the x-axis reflect the distribution of observed values. The p-value for the nonlinear component of each spline is shown in the upper left corner of each RCS panel.

Abbreviations: ARDS, acute respiratory distress syndrome; HR, hazard ratio; CI, confidence interval; RCS, restricted cubic spline.

**Supplementary Figure 10. Association Between Thoracic Body Composition Metrics and 60-Day Mortality in Patients with Acute Respiratory Distress Syndrome.**

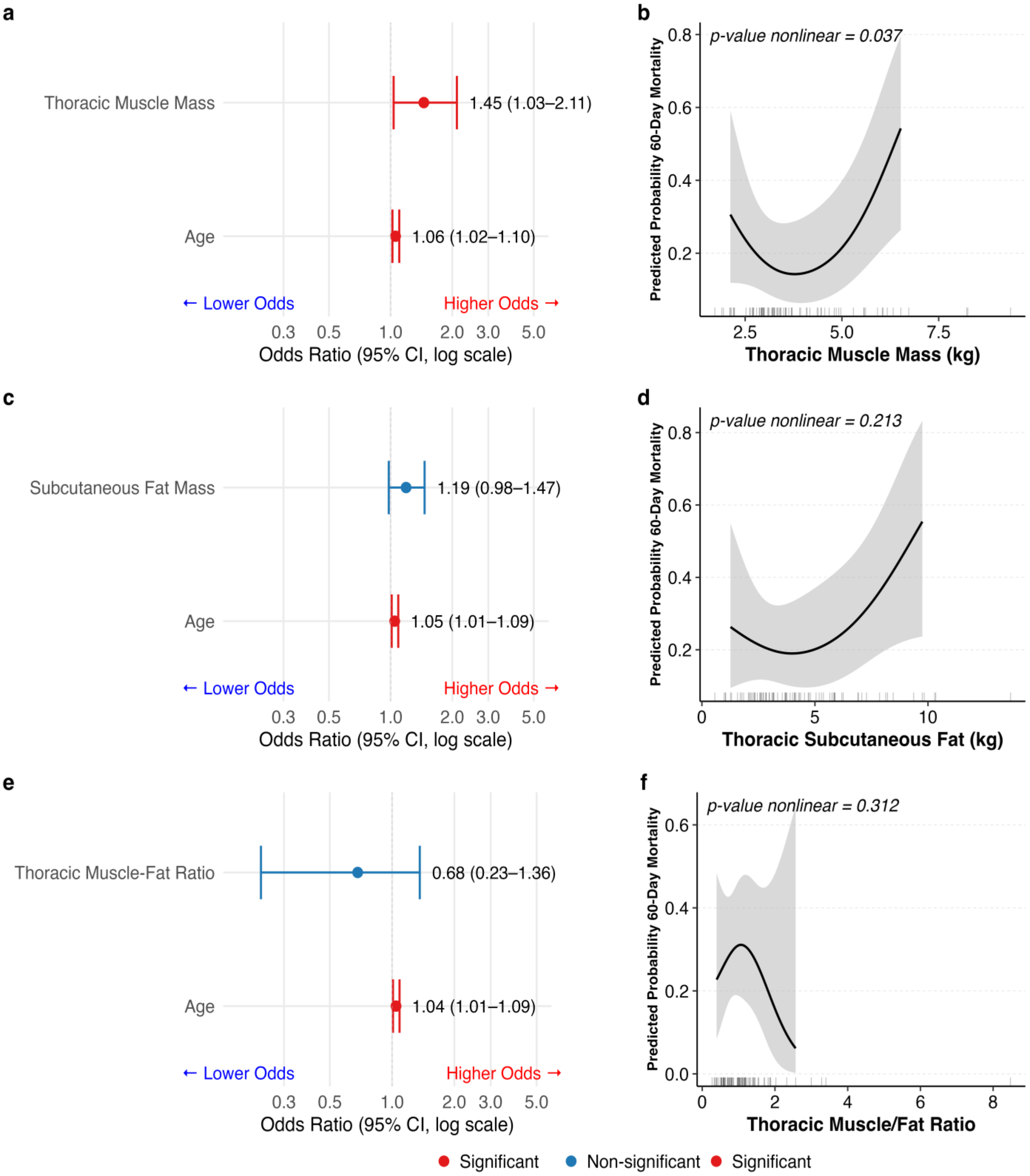

Panels (a), (c), and (e) display forest plots from multivariable logistic regression models examining the association of thoracic muscle mass (a), thoracic subcutaneous fat mass (c), and thoracic muscle-to-fat ratio (e) with 60-day mortality, adjusted for age in ARDS. Each point represents the odds ratio (OR) with 95% confidence intervals (CI); values greater than 1 indicate increased odds of mortality (risk factor) and values less than 1 indicate decreased odds of mortality (protective). Red points and error bars denote statistically significant associations ($p < 0.05$); blue denotes non-significant associations. Panels (b), (d), and (f) display restricted cubic spline (RCS) curves from corresponding logistic regression models, illustrating the dose-response relationship between each body composition metric and the odds of 60-day mortality. The shaded region represents the 95% CI. The dashed red horizontal line indicates an odds ratio of 1 (no effect). Rug marks along the x-axis reflect the distribution of observed values. The p-value for the nonlinear component of each spline is shown in the upper left corner of each RCS panel.

Abbreviations: ARDS, acute respiratory distress syndrome; OR, odds ratio; CI, confidence interval; RCS, restricted cubic spline.

**Supplementary Figure 11. Association Between BMI and 60-Day Ventilator Liberation and Mortality: Linear and Restricted Cubic Spline Models.**

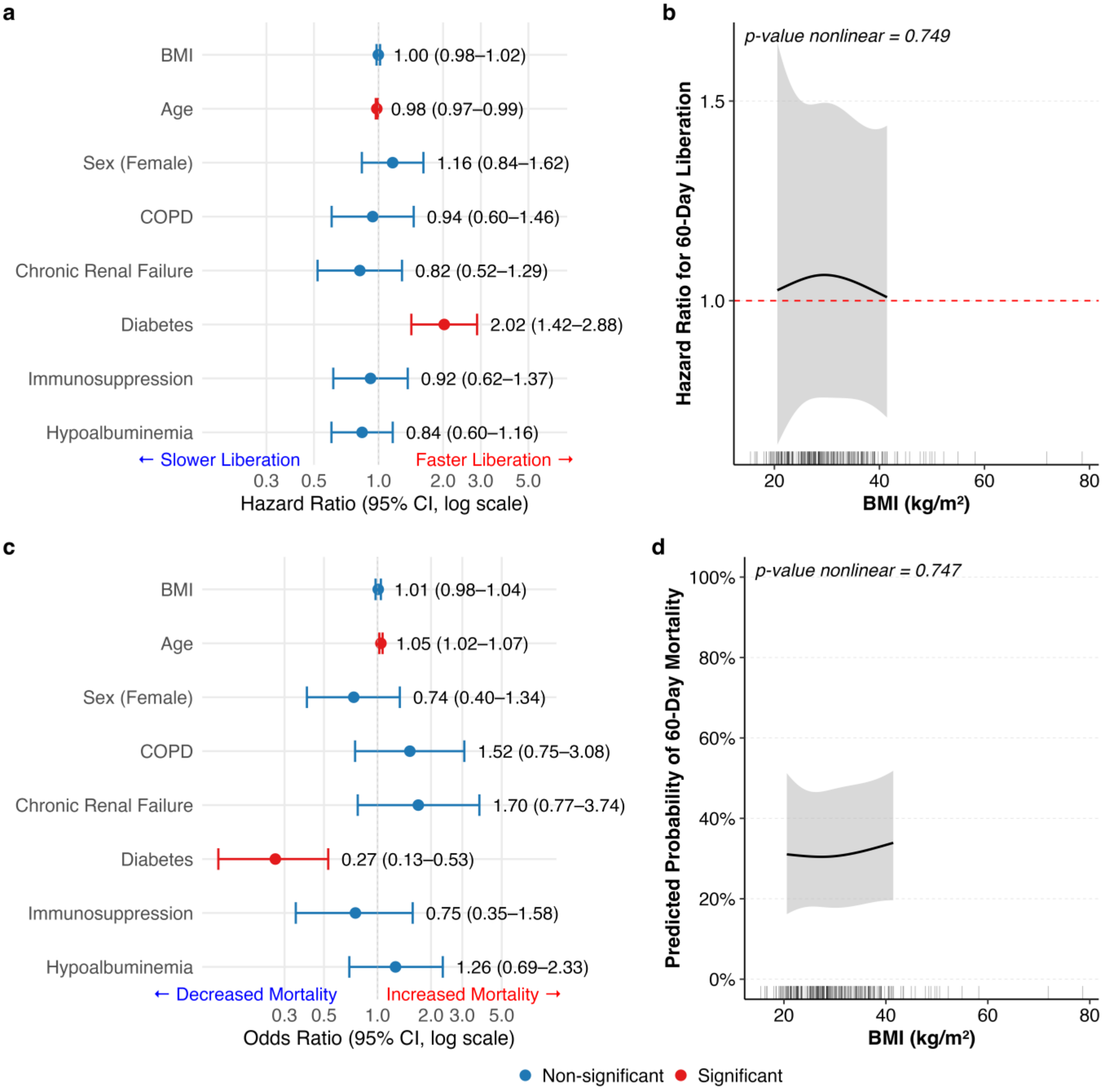


**(a)** Forest plot of a multivariable Cox model for 60-day ventilator liberation showing the hazard ratio (HR) for BMI modeled linearly, adjusted for age, sex, COPD, diabetes, chronic renal failure, immunosuppression, and hypoalbuminemia; red = significant ($p<0.05$), blue = non-significant. **(b)** Restricted cubic spline (RCS, 3 knots) plot of adjusted HR for liberation across BMI from the same model, with 95% CI (gray band) and nonlinearity p-value shown; dashed line marks HR = 1. **(c)** Forest plot of a multivariable logistic regression model for 60-day mortality showing the odds ratio (OR) for linear BMI, adjusted for the same covariates. **(d)** RCS plot of predicted probability of 60-day mortality across BMI from the same model, with 95% CI and nonlinearity p-value. Rug marks indicate observed BMI distribution.

Abbreviations: BMI = body mass index; CI = confidence interval; HR = hazard ratio; OR = odds ratio; RCS = restricted cubic spline.
